# Genetic predisposition to longer lifespan, lifestyle factors, and all-cause mortality: a 17-year prospective cohort study

**DOI:** 10.1101/2025.08.19.25333952

**Authors:** Niko Paavo Tynkkynen, Laura Joensuu, Päivi Herranen, Jaakko Kaprio, Timo Törmäkangas, Elina Sillanpää

## Abstract

**Background:** We used a genome-wide polygenic lifespan score (PLS) to investigate whether genetic predisposition to a longer lifespan is associated with lower mortality risk and how this association compares with associations between long-term lifestyle factors and mortality.

**Methods:** A PLS was computed for the Older Finnish Twin Cohort (mean age 57·4 years; 45·2% males; N=5575). Cox regression models were used to analyse the effects of the PLS on all-cause mortality risk before and after adding covariates (sex, physical activity, BMI, alcohol consumption, smoking, and education level). Differences between concordance indices (C-indices) were used to evaluate each predictor’s contribution to the model’s discriminatory performance.

**Findings:** Over a mean follow-up of 17·5±8·3 years, 1405 deaths (25·2%) were recorded. A one standard deviation increase in the PLS was statistically significantly associated with a lower all-cause mortality risk (hazard ratio [HR]=0·838, 95% confidence interval [CI]=0·792–0·887) and remained relatively unchanged after adding all covariates (HR=0·863, 95% CI=0·816–0·912). Smoking 20 or more cigarettes per day showed the strongest association with increased mortality risk (HR=3·341, 95% CI=2·751–4·056), whereas female sex was linked to the greatest risk reduction (HR=0·678, 95% CI=0·597–0·770). Smoking had the largest impact on model discrimination (ΔC-index=0·027), whereas all other covariates increased the C-index by less than 0·006.

**Interpretation:** Genetic predisposition to a longer lifespan was associated with a small decrease in all-cause mortality risk. This association remained after adjusting for lifestyle factors and covariates. Smoking behaviour and female sex were stronger predictors of mortality than genetic scores for lifespan.

**Funding:** This study was funded by the Research Council of Finland (341750, 346509, and 361981 for ES) and the Juho Vainio, and Päivikki and Sakari Sohlberg Foundation (ES). Finnish Twin Cohort data collection was supported by the Research Council of Finland (264146, 308248, 336823, and 352792 for JK), the Wellcome Trust Sanger Institute, the Broad Institute, ENGAGE, and FP7-HEALTH-F4-2007 (201413).

**Research in context:** *Evidence before this study:* We systematically searched PubMed and Google Scholar for studies related to the association of polygenic scores of lifespan/longevity and lifestyle factors with the risk of mortality from inception to November 30, 2024, and published in the English language. The search revealed three noteworthy publications on the effects of lifestyle factors and genetic predisposition to lifespan on mortality risk. However, the results regarding sex-specific results and information on the contribution of different factors in predicting deaths were scarce. While lifespan may be influenced by both genetic and lifestyle factors, it remains unclear how separate lifestyle factors affect the relationship between the genetic determinants of lifespan and mortality. Additionally, understanding how well genes and lifestyle factors predict all-cause mortality is crucial for evaluating their contribution to human lifespan.

*Added value of this study:* Our work presents results on the interplay between genetic predisposition to lifespan and lifestyle factors in all-cause mortality. We demonstrate the relative contribution of genetic and lifestyle factors and mortality-related demographics in predicting all-cause mortality. Furthermore, we provide sex-specific associations.

*Implications of all the available evidence:* Based on this study, the association between genetic predisposition to a longer lifespan and lower risk of all-cause mortality remains relatively independent of lifestyle factors and known covariates. However, female sex and smoking behaviour remain important, albeit opposing, factors in all-cause mortality. The findings of this study can be used to guide future research on lifespan and longevity in the field of genetics.

## Introduction

A long lifespan, or longevity, can be seen as a ‘health-positive’ trait, as it may indicate an adherence to healthy lifestyles^1^ and the enhanced avoidance of diseases.^2^ Human lifespan may be affected by genetic, lifestyle related, and environmental factors. Additionally, genetic variation can influence the lifestyle factors an individual adopts. For example, some individuals may inherit gene variants that make them naturally more inclined to follow a healthy diet^3^ and exercise routine,^4^ which may promote a longer lifespan. Healthy lifestyles, such as a balanced diet and regular exercise, can promote longevity e.g. by reducing the risk of chronic diseases.^5^ Genetic factors may protect against disease and promote longevity, but it remains unclear whether their overall impact outweighs that of lifestyles.

The average human lifespan is quite long, which makes it technically difficult to study how genetics affect lifespans. Moreover, as biobank infrastructures are relatively recent, most studies have limited follow-up durations, and only a small proportion of participants have reached the end of life. Consequently, datasets that combine genotype information with comprehensive lifestyle data and fully observed lifespans remain scarce. Trying to address the challenge of largely right-censored lifetime data, researchers have used the parental lifespan as a lifespan phenotype.^6^ However, this does not help in excluding the problem of left-censoring/truncation, which arises when follow-up begins mid-life, introducing potential selection bias due to the exclusion of individuals who died before enrolment. The association between parents’ lifespan phenotypes and descendants’ genotypes is assessed as an indirect association in genome-wide association studies (GWASs). Information from GWASs has been used to construct polygenic scores (PGSs), which have enabled researchers to study the effects of the whole genome on lifespan phenotypes.^6,7^ Only a few studies have investigated how lifestyle factors affect the association of genetic predisposition to lifespan and the risk of all-cause mortality.^8–10^ The evidence has limited sex-specific findings and information on the absolute effect sizes of genetic and lifestyle factors in explaining mortality risk.

In this study, we investigate if the genetic predisposition to a longer lifespan (polygenic lifespan score, PLS^6^) is associated with the risk of all-cause mortality and how adjustments for leisure-time physical activity (LTPA), body mass index (BMI), alcohol consumption, smoking behaviour, sex, and education level alter this association in the Older Finnish Twin Cohort. We also investigate the ability of these factors to predict the observed ranking of the participants with respect to their deaths from any cause. We hypothesise that the PLS is statistically significantly associated with a lower risk of all-cause mortality but that some lifestyle factors, especially smoking behaviour, may predict mortality better than the PLS.

## Methods

### Study design and participants

We constructed the PLS for a subsample of 8815 genotyped individuals in the Older Finnish Twin Cohort (FTC).^11^ Our final study sample, after excluding participants with missing data, included 5576 genotyped participants (mean age at DNA sampling: 57·4±10.0 years, 45·2% males). The FTC has four survey waves: baseline data collection in 1975 and follow-ups in 1981, 1990, and 2011–2012. The study sample was primarily defined based on data from 1990. Missing information was supplemented, when available, using data from 1981. The study received ethical approval from the University of Helsinki and Helsinki University Central Hospital ethical committees. All participants signed informed consent forms. Data collection followed the Declaration of Helsinki.

### Procedures

The PLS was constructed from the Older FTC sample using GWAS data from Timmers and colleagues^6^ The GWAS dataset included 1 012 240 parental lifespan phenotype measurements (UK Biobank: 339 732 paternal and 351 889 maternal, LifeGen consortium: 160 461 paternal and 160 158 maternal) and provided summary statistics based on 500 193 European individuals (offspring) and for 9 085 648 genetic variants. The UK Biobank study was approved by the North West Multi-Centre Research Ethics Committee (approval no. 11/NW/0382). LifeGen consortium cohort studies were approved by their respective local committees, with written informed consent obtained from each participant. Information on the genotyping, quality control, and imputation of the Older FTC samples and the cohorts used in the GWAS (the LifeGen consortium and UK Biobank) are shown in the appendix (p. 3). The PLS was computed using the SBayesR methodology.^12^ The parental lifespan GWAS summary statistics were restricted to 1 180 172 common genetic variants (MAF>5%) of the HapMap3 variant list.^13^ Using common variants as the basis for a genetic score has demonstrated good transferability across different populations.^14^ PLINK v1.9 (version 1.90b6.9 64-bit) software was used to calculate the PLS based on 992 782 genetic variants. A higher genetic score indicated a genetic predisposition to a longer lifespan.

### Outcomes

All-cause mortality outcomes were based on death records from the Population Register Centre of Finland and Statistics Finland. These data were linked to our study sample’s genetic and lifestyle information using personal identity codes assigned to every permanent Finnish resident. Survival time was calculated as the age interval between personal follow-up entry (age at DNA sampling) and end age. Age was computed from birth (“origin”), but follow-up started from the date of DNA collection. The DNA samples were collected between 1993 and 2017. End age was defined as the date of emigration, date of death, or end of follow-up (December 31, 2020).

### Covariates

Covariates sex (male or female), LTPA (MET-h/wk), BMI (kg/m^2^), alcohol consumption (g/d), and smoking behaviour (never-smokers, occasional, former, light [1–9 cigarettes per day, CPD], medium [10–19 CPD], and heavy smokers [20 or more CPD]), and education level (years of education) were selected based on previous research and availability.^15–19^ LTPA was constructed as an average of three surveys (1975, 1981, and 1990). BMI was calculated from self-reported weight and height in 1981. Alcohol consumption and smoking behaviour were determined based on data from 1990. If information was missing, it was supplemented, when possible, using data from 1981. Genetic stratification was adjusted using the first ten principal components (PCs) of Finnish ancestry. Family relatedness (family identifiers) was included as a random effect to account for twin-pair relatedness. Details of the covariates conducted with the Older FTC data are presented in the appendix (pp. 3–4).

### Statistical analysis

We analysed the association between the risk of all-cause mortality and the PLS using mixed-effects Cox proportional hazards models computed using the coxme function in R software’s *coxme* package (version 2.2-22).^20^ Mortality risk was calculated per one standard deviation (SD) change in the PLS, with the results reported as hazard ratios (HRs) and 95% confidence intervals (CIs). The first model included PLS as the predictor and was adjusted for the first ten genetic PCs and family relatedness.

Kaplan–Meier analysis was used to describe and assess how survival differs based on one’s PLS by comparing survival between PLSs grouped as low (first tertile), middle (second tertile), and high (third tertile). Log-rank testing was used to compare survival between the groups and pairwise by comparing the lowest and highest groups. Due to left truncation in our data, log-rank test results were obtained from bivariate Cox regression models using the coxph function of the *survival* package (version 2.2-22).^21^ Survival graphs were created using the *ggsurvfit* package (version 1.1.0)^22^ and the median survival times were calculated using the survfit function of the *survival* package (version 2.2-22).^21^

In the subsequent mixed-effects Cox proportional hazards models, sex, LTPA, BMI, alcohol consumption, and smoking behaviour were added individually as covariates (Models 2–6, respectively), while Model 7 included all lifestyle factors. Model 8 accounted for education level separately and Model 9 incorporated all covariates. Proportional hazards assumption tests and visual inspections showed no major violations of the proportional hazards assumption (see the appendix pp. 11 and 20–31). While light and medium smoking statistically violated the proportional hazards assumption in the full sample, visual inspection of the Schoenfeld residuals did not indicate meaningful violations. The proportional hazards assumption violation of the PCs was neglected, as the PCs were considered adjusting factors. No notable outliers were detected based on Martingale and deviance residuals, and the linearity assumption was met. We tested interactions between the PLS, sex, lifestyle factors, and education level. Statistical significance was set at a *p*-value of less than 0·05.

To evaluate the individual contribution of each covariate (PLS, sex, lifestyle factors, and education level) to mortality, we calculated the difference in concordance indices (ΔC-indices) between the full model (including all covariates and adjustments) and a reduced model in which the covariate of interest was omitted. The C-indices were derived from complete case Cox regression models. All models were adjusted for the first ten genetic PCs of ancestry. To calculate the ΔC-indices for the individual smoking behaviour categories, five binary smoking behaviour variables were constructed out of the original categorical smoking behaviour variable (six categories). The group of never-smokers was the reference group.

Additionally, we investigated whether the PLS has *indirect effects* through sex, lifestyle factors, and/or education level. We analysed the associations of the PLS with all these factors using bivariate linear mixed-effects models in the complete case dataset (N=5411). All continuous variables were standardized prior to analysis. The models were constructed using the *nmle* package (version 3.1-166)^23^ in R software. Family relatedness was included in the model as a random intercept.

We performed several *sensitivity analyses*. Sex-specific analyses included a Cox regression analysis, C-index calculations, and Kaplan–Meier analysis for males and females separately. We performed an additional Cox regression analysis to account for varying levels of missing data in the variables (N=9–165), using complete case data for all study variables in all participants, males, and females. Finally, we performed a Cox regression analysis including all other factors expect the PLS and the first ten genetic PCs of ancestry. The analysis protocol followed in the sensitivity analyses was similar to that followed in the main analyses.

## Results

Baseline characteristics are presented in Table 1. From the start of the first DNA sampling in 1993 to the end of the mortality follow-up in 2020, there were 1405 deaths (25·20%) in 5576 participants. The mean follow-up time was 17·50±8·32 years (see the histogram in the appendix p. 17). In general, the participants had quite low levels of physical activity, had a median of 7 years of education, were slightly overweight, consumed on average 9·56 grams of alcohol per day, and were mostly (72·76%) non-smokers (never or former smokers). Details of the distribution of participants’ PLS values can be found in the appendix (p. 17).

**Table 1.** Descriptive statistics of the Finnish Twin Cohort study data. The numbers of deaths were reported from the models, including the highest number of participants. The polygenic lifespan score (PLS) was z-standardized. SD=standard deviation. IQR=Interquartile range. n=number of participants in a category. N=total number of participants. LTPA=leisure-time physical activity. MET=metabolic equivalent of task. CPD=cigarettes per day.

|  | Mean (SD) | IQR | N |
| --- | --- | --- | --- |
| Age at the follow-up entry (years) | 57.44 (9.99) | 13.95 | 5576 |
| Age at the follow-up end (years) | 74.94 (7.88) | 12.46 | 5576 |
| PLS (per 10<sup>7</sup> units) | -0.23 (0.95) | 1.30 | 5576 |
| LTPA (MET-h/wk) | 2.64 (2.22) | 2.17 | 5576 |
| BMI (kg/m<sup>2</sup>) | 25.00 (3.80) | 4.62 | 5573 |
| Alcohol consumption (g/d) | 9.56 (14.17) | 9.03 | 5573 |
| Education level (years) | 8.15 (2.97) | 4.00 | 5420 |
|  | n | % | N |
| **Number of participants** |  |  |  |
| Male | 2522 | 45.23 |  |
| Female | 3054 | 54.77 |  |
| **Number of deaths** |  |  | 1405 |
| Male | 807 | 57.44 |  |
| Female | 598 | 42.56 |  |
| **Smoking behaviour** |  |  | 5573 |
| Never | 2668 | 47.87 |  |
| Occasional | 169 | 3.03 |  |
| Former | 1387 | 24.89 |  |
| Light (1-9 CPD) | 297 | 5.33 |  |
| Medium (10-19 CPD) | 559 | 10.03 |  |
| Heavy (20 or more CPD) | 493 | 8.84 |  |

The Kaplan–Meier graph in Figure 1 shows that the median ages of survival were 83·70 years (95% CI=82·98–84·36) for the low, 86·22 years (85·38–87·12) for the middle, and 86·97 years (86·01–88·13) for the high PLS group. Survival differed statistically significantly between all groups (log-rank χ^2^[degrees of freedom=2]=39·43, *p*=2·75×10^−9^). Pairwise comparisons showed statistically significant survival differences between the low and middle PLS groups (*χ*^2^[1]=16·03, Bonferroni-adjusted *p*=1·87×10^−4^), and between the low and high (*χ*^2^[1]=37·06, Bonferroni-adjusted *p*=3·44×10^−9^) PLS groups. The median survival age in the high PLS group exceeded that of the low and middle groups by 0·76 to 3·28 years.

**Figure 1.**
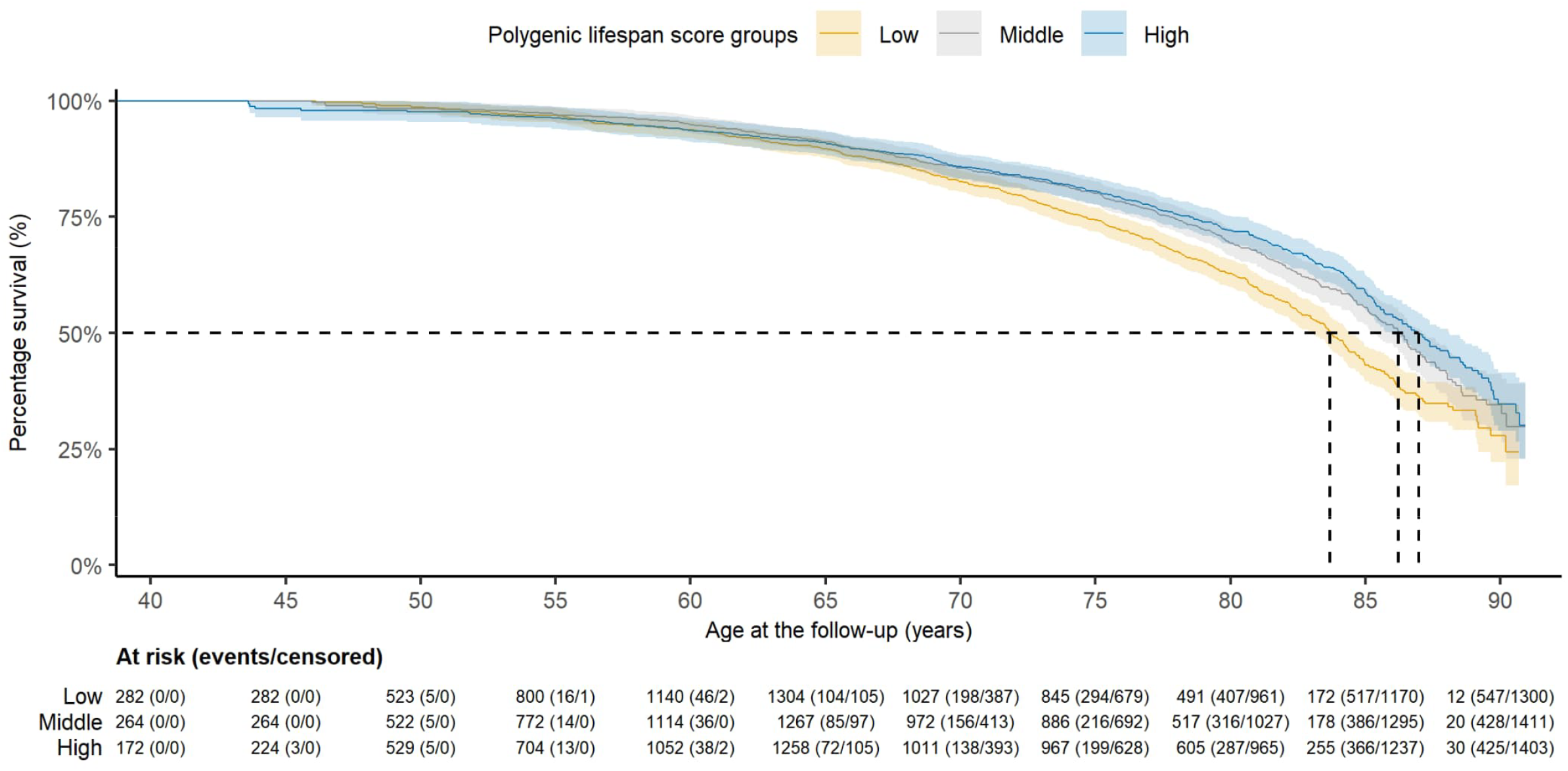
Kaplan–Meier survival curves for all-cause mortality risk in the three PLS groups. Median survival is indicated using a dashed line.

One SD increase in the PLS was statistically significantly associated with a lower risk of all-cause mortality (HR=0·838, 95% *CI*=0·792–0·887, ΔC-index=0·005; Model 1, Table 2). The association between PLS and mortality remained statistically significant across all adjusted models (highest, model 9, HR=0·863, 95% CI=0·816–0·912).

**Table 2.** Associations of the PLS and lifestyle factors with the risk of all-cause mortality in the Older Finnish Twin Cohort. The table includes the hazard ratios and their 95% confidence intervals for each covariate inspected in the different models. PLS=polygenic lifespan score. LTPA=leisure-time physical activity. BMI=body mass index. Model 1=all-cause mortality ~ PLS + the first ten genetic principal components of ancestry + family relatedness as random factor. Model 2=Model 1 + sex. Model 3=Model 1 + LTPA. Model 4=Model 1 + BMI. Model 5=Model 1 + alcohol consumption. Model 6=Model 1 + smoking behaviour. Model 7=Model 1 + lifestyle covariates. Model 8=Model 1 + education level. Model 9=Model 1 + all covariates. Bold type indicates statistical significance at the level of *p*<0·05.

| Deaths/Participants | Model 1<br>1405/5576 | Model 2<br>1405/5576 | Model 3<br>1405/5576 | Model 4<br>1404/5573 | Model 5<br>1404/5573 | Model 6<br>1404/5573 | Model 7<br>1402/5567 | Model 8<br>1368/5420 | Model 9<br>1365/5411 |
| --- | --- | --- | --- | --- | --- | --- | --- | --- | --- |
| PLS | **0·838**<br>(0·792–0·887) | **0·834**<br>(0·789–0·881) | **0·838**<br>(0·792–0·887) | **0·845**<br>(0·799–0·894) | **0·837**<br>(0·791–0·886) | **0·859**<br>(0·813–0·908) | **0·862**<br>(0·815–0·911) | **0·845**<br>(0·798–0·895) | **0·863**<br>(0·816–0·912) |
| Sex |  | **0·498**<br>(0·444–0·559) |  |  |  |  |  |  | **0·678**<br>(0·597–0·770) |
| LTPA<br>(MET-h/week) |  |  | **0·959**<br>(0·932–0·987) |  |  |  | **0·971**<br>(0·944–0·998) |  | **0·968**<br>(0·941–0·995) |
| BMI<br>(kg/m<sup>2</sup>) |  |  |  | **1·040**<br>(1·025–1·055) |  |  | **1·037**<br>(1·022–1·052) |  | **1·031**<br>(1·015–1·047) |
| Alcohol consumption<br>(g/d) |  |  |  |  | **1·021**<br>(1·018–1·024) |  | **1·013**<br>(1·010–1·016) |  | **1·011**<br>(1·007–1·014) |
| Smoking behaviour |  |  |  |  |  |  |  |  |  |
| Never |  |  |  |  |  | 1·000 (ref.) | 1·000 (ref.) |  | 1·000 (ref.) |
| Occasional |  |  |  |  |  | **1·465**<br>(1·044–2·054) | 1·316<br>(0·938–1·848) |  | 1·278<br>(0·907–1·801) |
| Former |  |  |  |  |  | **1·647**<br>(1·429–1·898) | **1·445**<br>(1·249–1·671) |  | **1·277**<br>(1·097–1·488) |
| Light |  |  |  |  |  | **1·803**<br>(1·375–2·363) | **1·750**<br>(1·334–2·294) |  | **1·666**<br>(1·267–2·189) |
| Medium |  |  |  |  |  | **2·445**<br>(2·023–2·954) | **2·268**<br>(1·873–2·746) |  | **2·078**<br>(1·713–2·522) |
| Heavy |  |  |  |  |  | **4·787**<br>(4·010–5·715) | **3·764**<br>(3·115–4·549) |  | **3·341**<br>(2·751–4·056) |
| Education level |  |  |  |  |  |  |  | **0·960**<br>(0·938–0·982) | **0·970**<br>(0·948–0·991) |

Sex, all lifestyle factors, and education level had statistically significant associations with the risk of all-cause mortality conditional on the PLS in all models (Models 2–9, Table 2). Being female was associated with the greatest risk reduction in all-cause mortality (lowest HR=0·498, 95% CI=0·444–0·559; highest HR=0·678, 95% CI=0·597–0·770; models 2, 7, and 9, Table 2). The risk of all-cause mortality increased with the intensity of smoking when the never-smoking group was used as the reference group. Heavy smoking showed the strongest association with increased all-cause mortality risk (lowest HR=3·341, 95% CI=2·751–4·056; highest HR=4·787, 95% CI=4·010–5·715). Of the tested covariates, sex and smoking behaviour contributed the most to the model’s discriminatory ability to predict deaths, as shown by the largest reductions in the C-index when these variables were omitted (ΔC-index 0·006 and 0·027; Table 3). The contributions of other lifestyle factors and education levels on the model’s discriminatory ability in predicting all-cause mortality were limited.

**Table 3.** Concordance index for the models omitting each covariate in turn from the full model, and differences between the full model and the omitted-covariate models. C-index=concordance index. SE=standard error. ΔC-index=difference in C-index between the full model and a model without a covariate. PLS=polygenic lifespan score. LTPA=leisure-time physical activity. BMI=body mass index. The C-index for the model (full model) including all covariates and the first ten genetic principal components of ancestry was 0·664 (SE=0·008). The number of deaths was 1365 and the number of participants was 5411.

| | C-index without the variable (SE) | ΔC-index |
| --- | --- | --- |
| **PLS** | 0·659 (0·008) | 0·005 |
| **Sex** | 0·658 (0·008) | 0·006 |
| **LTPA** | 0·663 (0·008) | 0·001 |
| **BMI** | 0·660 (0·008) | 0·004 |
| **Alcohol consumption** | 0·659 (0·008) | 0·005 |
| **Smoking behaviour** | 0·637 (0·009) | 0·027 |
| Never | ref. | 0 |
| Occasional | 0·664 (0·008) | 0·000 |
| Former | 0·662 (0·008) | 0·002 |
| Light | 0·661 (0·008) | 0·003 |
| Medium | 0·653 (0·009) | 0·011 |
| Heavy | 0·643 (0·008) | 0·021 |
| **Education level** | 0·663 (0·008) | 0·001 |

The PLS did not have statistically significant interactions with sex; LTPA; BMI; alcohol consumption; occasional, former, light, or heavy smoking; or education level (*p*>0·05). However, it had a statistically significant interaction with medium smoking (see supplementary appendix, p. 11). We suggest that this single interaction in a subset can most likely explained by the uneven distribution of the participants in the smoking behaviour groups and by not conducting further modelling including an interaction term.

In the linear and non-linear mixed-effects modelling analysis, the standardized regression coefficients (beta) ranged from −0·108 to 0·085 for associations between the PLS and other fixed-effects covariates, indicating minimal indirect effects (appendix p. 9). Even though we did not observe a statistically significant PLS×sex interaction, we provide Cox regression, ΔC-index, and Kaplan–Meier results for males and females, which are presented in the appendix (pp. 6–8, 10, and 18–19). Overall, the sex-specific analyses revealed rather similar associations among males and females compared to what was observed in the main analysis. Complete case analyses showed Cox regression results (appendix pp. 12–15) like the main analysis, indicating that missing covariate observations did not affect the results. The analysis excluding the PLS and the first ten genetic PCs of ancestry showed very minimalistic changes (appendix p. 16).

## Discussion

We examined whether the PLS is associated with all-cause mortality risk and how sex, lifestyle factors (LTPA, BMI, alcohol use, smoking), and education influence this association. A higher genetic predisposition to longer lifespan associated with lower mortality risk, and this association remained largely unchanged after adjusting for covariates. Sex and smoking had the strongest independent effects. Overall, the PLS and covariates showed low discrimination ability, consistent with prior evidence that lifespan is modestly heritable and PLS explains only a small portion of its variation.

The genetic basis of lifespan is complex. While lifestyle and environment strongly influence longevity, genetics also contribute. Prior studies link variants in genes like *APOE* and *FOXO3* to increased lifespan.^24^ Our findings support a weak association between genetic liability and longer lifespan, possibly via effects on metabolism, cellular repair, and disease resistance. Although sex interactions were not statistically significant, the PLS showed slightly stronger protective effects in males. This may reflect that common variants offer greater protection to males, while females may benefit from additional mechanisms like asymmetric inheritance of sex chromosomes and mitochondria.^25^ Differences in survival between sexes may also stem from random variation, other lifestyle factors and environmental exposures, or differing reasons for DNA sample submission. Comparing sexes is further complicated by higher mortality risk in males, with over 10 percentage points more deaths than females. Further, the increase in life-expectancy in Finland has been substantial in recent time, being slightly over 60 years for males immediately after WWII, compared to nearly 80 years at present. The difference in life-expectancy between males and females has decreased in the past decades, attributable primarily to strong declines in smoking among Finnish males.

Lifespan is shaped by a complex interaction between genetics and lifestyle. While genetic predispositions influence health outcomes, lifestyle and environmental factors play a major role in their manifestation. Wang and colleagues^8^ examined whether a genetic score based on 11 SNPs interacts with a lifestyle score (four factors) in predicting mortality. Healthy lifestyles were linked to lower mortality, especially among those with high genetic risk, though differences between genetic risk groups were small and statistically non-significant. Bian and colleagues^9^ and Li and colleagues^10^ used similar designs, with Li’s and colleagues’ polygenic score based on 331 SNPs showing that lifestyle had a stronger impact on lifespan than genetic predisposition, which aligns with our findings. All three studies found healthier lifestyles associated with reduced mortality risk, but limited evidence that high-risk individuals benefit more. Most prior research has emphasised composite lifestyle scores, with limited focus on specific factors,^8–10^ though specific factors like device-measured physical activity and sedentary behaviour may show similar associations.^26^ Although effect sizes were not consistently reported, hazard ratios suggest small effects, in line with our results.

We provide novel evidence that the PLS is associated with the risk of all-cause mortality, independent of lifestyle factors and mortality-related demographics. These findings align with a previous twin study suggesting that monozygotic twins resemble each other more than dizygotic twins do in terms of longevity^27^, although the twin study implies higher heritability for longevity than found in GWAS studies. However, it is noteworthy that twin studies typically report modest heritability estimates for lifespan. This is consistent with findings across species, where heritability estimates for lifespan tend to be low. Based on the ΔC-indices, the PLS had lower predictive ability than smoking behaviour and sex but was comparable to or exceeded alcohol consumption, BMI, physical activity, and educational level.

Our study has several strengths. First, the Older FTC provided extensive, well-validated questionnaire-based lifestyle and socioeconomic data across multiple time points. Second, the Finnish causes of death registry is a high-quality source of comprehensive and accurate mortality data meticulously compiled from the Population Information System and death certificates. Maintained since 1936, the registry is renowned for its reliability and completeness, making it an invaluable resource for epidemiological research.^28^ Third, the GWAS summary statistic used as base data for the PLS computation was based on a very large number of participants. A larger number of GWAS participants yielded more accurate genetic associations, which, in turn, provided more reliable weights for the PLS computation. Fourth, our study explored time-to-event relationships in extensive survival analyses, with validating model assumption inspections and supporting sensitivity analyses. Fifth, our analyses added sex-specific knowledge of how genetic predisposition to lifespan is associated with all-cause mortality and how this association is affected by lifestyle factors. Lastly, analysing how well the study’s factors predicted the observed ranking of deaths using C-indices provided insights into the relative predictive value of each factor, particularly the PLS.

The results should be interpreted with the following limitations in mind. First, the adapted PLS is an indirect lifespan estimator derived from GWAS data on genetic associations between parental lifespan and offspring genotypes. Parents and offspring may have experienced different environmental exposures that interacted with lifespan-related genes, which may have affected the accuracy and relevance of genetic association estimates. Lifespan phenotyping requires comprehensive life course data, which large-scale biobanks currently lack. Second, some lifestyle factors were averaged across three time points, possibly leading to over- or underestimation. LTPA is especially prone to this bias due to the variability in individual behaviour over time. Also, using GWAS data from participants with ancestries different from those in the PLS computation may have introduced bias.^29^ The Finnish population has some genetic differences compared to the British and other European ancestries used in the base GWAS. Third, genes located on sex chromosomes were not included in the analyses, which may have led to the omission of sex-specific genetic effects on lifespan.^30^ Lastly, the small effect size of the PLS may also be related to the possibility that the lifespan is mainly driven by non-genetic effects, and we only utilized a limited set of environmental exposures to assess these effects.

In conclusion, genetic predisposition to a longer lifespan was associated with a decreased risk of all-cause mortality, albeit with a small effect size. This association remained unchanged after adjusting for lifestyle factors and other known covariates. Of all the evaluated variables, smoking behaviour and female sex were the most important contributors to all-cause mortality, with smoking increasing and female sex decreasing the risk. Based on the evidence of this study, lifestyle, social, and environmental factors may have stronger and more substantial effects on lifespan. However, the small effect size suggests that PLS has little clinical utility. To better understand how lifestyle factors interact with genetics, future research should focus on developing more precisely designed GWASs for lifespan and longevity.

## Contributors

ES conceived of the idea for the study. ES designed and supervised the construction of the PLS, and NPT conducted the analyses under the supervision of TP. LJ assisted with data handling and prerequisites. NPT designed and ES and TT supervised the statistical analysis, with PH’s assistance. NPT performed the statistical modelling. NPT, ES, TT, JK and PH performed the interpretation of the data. NPT drafted the first version of the manuscript, and ES, LJ, PH, JK and TT contributed significantly to the writing. ES and JK acquired the funding for this study, and JK also acquired funding for the Older FTC data collection and genotyping. All authors were involved in drafting the manuscript and revising it critically for important intellectual content. They approve the analyses conducted and provide their final approval of this version to be published.

## Data sharing

The GWAS summary statistics used are publicly available online via the Edinburgh DataShare web page (https://datashare.ed.ac.uk/handle/10283/3209). The Older FTC data are not publicly available, but authorised researchers can apply for access via the THL Biobank, provided they have institutional review board/ethical approval and an institutionally approved study plan. For details, see the THL Biobank website (https://thl.fi/en/research-and-development/thl-biobank/for-researchers).

## Acknowledgements

The authors thank all the FTC study participants, the FTC research team members, and the persons involved in the data collection. The Gerontology Research Centre is a joint effort between the University of Jyväskylä and the University of Tampere.

## Supplementary material

Supplementary appendix

