## Supplementary appendix for "Genetic predisposition to longer lifespan, lifestyle factors, and all-cause mortality: a 17-year prospective cohort study"

### Genotyping, quality control, and imputation of the study cohorts

#### The Older Finnish Twin Cohort

In the Older Finnish Twin Cohort (FTC), genotyping was conducted at three locations: the Human670‐QuadCustom Illumina BeadChip (Illumina, Inc., San Diego, CA, USA) at the Wellcome Trust Sanger Institute and the Illumina Human Core Exome BeadChip (Illumina) at the Wellcome Trust Sanger Institute and at the Broad Institute of MIT and Harvard. The exclusion criteria before imputation for the data generated using the Human670‐QuadCustom Illumina BeadChip included a minor allele frequency (MAF) of less than 0·01, a sample call rate of less than 0·98, and a single nucleotide polymorphism (SNP) call rate of less than 0·95 (<0·99 for SNPs with MAF<0·05). As for the data generated with the Illumina Human Core Exome BeadChip, the exclusion criteria were a minor allele count of less than 2, a sample call rate of less than 0·98, and an SNP call rate of less than 0·95 (<0·99 for SNPs with minor allele frequency<0·05). Both genotype datasets were filtered according to the Hardy–Weinberg equilibrium test (*p*<1×10^-6^). Outlier checks regarding the sample heterozygosity test, gender, and multidimensional scaling were conducted for the datasets. A detailed description of the genotyping, quality control, and imputation for the Older FTC study is provided in the studies of He and colleagues^1^ and Hällfors and colleagues.^2^

#### GWAS datasets: UK Biobank and LifeGen consortium cohorts

Genotyping of the UK Biobank samples was performed using the UK Biobank Axiom Array. Bycroft and colleagues^3^ described the processes conducted regarding genotyping, quality control, and imputation for the UK Biobank Study. The LifeGen consortium data consist of a collection of 26 datasets consisting of European and African ancestry datasets. The parental lifespan GWAS summary statistics used in this study included only European ancestry samples and detailed genotyping, quality control, and imputation. Related information is available in Joshi and colleagues^4^ (LifeGen consortium).

### Detailed descriptions of the education level, lifestyle factors, and the genetic principal components

#### Leisure-time physical activity

In 1975, 1981, and 1990, leisure-time physical activity (LTPA) questionnaires included questions regarding monthly frequency, session duration, and activity intensity. ^5,6^ In 1975 and 1981, information on the average intensity of LTPA was collected using the question:: ”*Is your leisure-time physical exercise, on average, as intensive as…*”, with response options: *“walking”*, *“walking and jogging”*, *“jogging”*, and *“running”*.^7^ These responses were assigned metabolic equivalent of task (MET) values as follows: 4 METs, 6 METs, 10 METs, and 13 METs.^8^ Information on the average duration of a single LTPA session was collected with the question: “*On average, how long does a single LTPA session last?*”, with response options: “*under 15 min*”, “*15 min – under half an hour*”, “*half an hour – less than an hour*”, “*an hour – less than two hours*”, and “*over two hours*”. These were coded as: 7.5 min, 22.5 min, 45 min, 90 min, and 120 min, respectively. Monthly frequency of LTPA was assessed with the question: “*How many times per month do you currently engage in leisure-time physical activity*?”, with response options: “*less than once per month*”, “*1–2 times per month*”, “*3–5 times per month*”, “*6–10 times per month*”, “*11–19 times per month*”, and “*over 20 times per month*”. These were coded as: 0.5, 1.5, 4, 8, 15, and 20 times. Daily MET-hours (MET-h/d) were calculated by multiplying monthly frequency, session duration, and intensity. Weekly MET-hours (MET-h/week) were derived by multiplying daily MET-hours by seven.

In 1990, information on the intensity, duration, and frequency of LTPA was collected differently than in 1975 and 1981. Participants were asked to report their average hours of physical activity (PA) during leisure time or commuting over the past 12 months. They were asked to indicate how many hours (“*none*”, “*in total less than ½ hour per week*”, “*in total ½–1 hour per week*”, “*in total 2–3 hours per week*”, and “*in total 4 hours or more per week*”) they had engaged in PA at each intensity level (“*walking*”, “*brisk walking*”, “*jogging*”, and “*running*”). These duration responses were coded as: 0 hours, 0.25 hours, 0.75 hours, 2.5 hours, and 4 hours. Intensity levels were coded according to the same MET values used in the 1975 and 1981 questionnaires. Weekly MET-hours were calculated by multiplying the duration of PA at each intensity level by its respective MET value and summing the MET-hours across all intensity levels.^6^

To obtain the average LTPA, the values from the three time points were summed and divided by three. If there was missing information in any of the time points, the LTPA value(s) were neglected in the calculation of the average LTPA.

#### Body mass index

Body mass index (BMI, kg/m^2^) was calculated based on self-reported height and weight. The last known BMI, calculated from the self-reported weight and height in 1981, was used in the analysis.^9^ If information was missing in 1981, it was complemented with information from 1990.

#### Alcohol consumption

Alcohol consumption was estimated based on separate questions regarding average weekly quantities of beer and/or wine, and average monthly quantities of spirits. We used the most recent available values from either 1981 or 1990. The quantities were determined for each type of alcoholic beverage using seven-point scales: the upper limit for beer was 48 bottles or more per week, for wine 10 bottles or more per week, and for spirits 20 bottles or more per month. Alcohol consumption was calculated by converting the reported volumes of each beverage type into grams of pure alcohol, which were then summed and converted into an estimate of grams per day (g/day). ^10^

#### Smoking behaviour

All participants were asked the following question: ‘Have you ever smoked more than 5–10 packs of cigarettes during your entire life?’ Participants who answered this question negatively were classified as never smokers. Participants who answered positively were asked a further question: ‘Do you smoke, or have you smoked cigarettes regularly – say daily, or almost daily – during your lifetime?’ Former smokers were classified as those who smoked regularly, daily, or almost daily before quitting. Occasional smokers were classified as those with irregular smoking patterns. Participants who smoked daily or almost daily were classified as current smokers and were further asked about the daily number of cigarettes they smoked. The original response options were as follows: (a) none, (b) less than 5, (c) 5–9, (d) 10–14, (e) 15–19, (f) 20–24, (g) 25–39, and (h) more than 40 cigarettes per day (CPD). We reclassified these responses as light smokers (<10 CPDs), medium smokers (10–19 CPDs), and heavy smokers (≥20 CPDs). We used the last known information from either 1981 or 1990.^11^

#### Education level

Education level was assessed based on the number of years of education collected through self-reported answers. The options were as follows: less than primary school (mean 3 years), primary school (6 years), at least 1 year of education beyond primary school (mean of 7 years), junior high school (9 years), at least 1 year of education beyond junior high school (mean of 10 years), high school graduate (12 years), at least 1 year of education beyond high school graduation (mean of 13), and university degree (16 years of education). The answers were collected in 1975 and 1981.^12^

#### Genetic principal components of ancestry

Genetic principal components (PCs) of ancestry were computed by obtaining a variance-standardised relationship matrix using autosomal single nucleotide variants over all chip genotyping batches consisting of multiple arrays. We included the first ten genetic PCs of ancestry as covariates to reduce the risk of false positives caused by population stratification.^13^ All single nucleotide variants were first merged into one dataset, excluding variants with a minor allele frequency of less than 5%, a missingness rate of more than 5%, and a Hardy–Weinberg test statistic *p*-value of less than 1×10^-6^. In the final phase, the set of variants was pruned to have low linkage disequilibrium (*r*^2^<0·2, with the variant count window set to 50), resulting in 10,485 remaining variants for PC analysis, conducted with PLINK v1.9^14,15^ using the–pca flag.

### Supplementary results

#### Sex-specific descriptive characteristics

The mean follow-up time was 16·59±8·18 years for 2522 males (807 deaths, 32·00% of males) and 18·26±8·36 years for 3054 females (598 deaths, 19·58% of females). Males and females had similar BMI and educational levels, but males were slightly more physically active (2·84 MET-h/wk vs. 2·47 MET-h/wk), consumed more alcohol (14·60 g/d vs. 5·39 g/d), and were more often former (34·54% vs. 16·93%) and current smokers (33·03% vs. 22·46%; Table S1).

#### Sex-specific Kaplan–Meier analysis by polygenic lifespan group

The Kaplan–Meier survival curves for males in Figure S1 show that the median ages of survival were 79·59 years (95% confidence interval [CI]=78·18–82·36) for the low, 84·12 years (82·36–83·94) for the middle, and 85·01 years (83·94–86·01) for the high polygenic lifespan score (PLS) group. Survival differed statistically significantly between all PLS groups (*χ*²[degrees of freedom=2]=46·68, *p*=7·29×10⁻^11^). Pairwise comparisons showed statistically significant differences in survival between the low and middle PLS groups (*χ*²[1]=24·75, Bonferroni-adjusted *p*=1·96×10⁻^6^), and between the low and high (*χ*²[1]=39·33, Bonferroni-adjusted *p*=1·07×10⁻^9^) PLS groups. The median survival age in the high PLS group exceeded that of the low and middle PLS groups by 0·89 to 5·43 years.

The Kaplan–Meier survival curves for females in Figure S2 show that the median survival ages were 87·00 years (86·08–89·65) for the low and 87·59 years (86·55–89·11) for the middle, and 89·56 (88·06–NA) for the high PLS group. Survival differed statistically significantly among all groups (*χ*²[2]=6·68, *p*=0·035). There was a statistically significant survival difference between the low and the high PLS groups (*χ*²[1]=5·85, Bonferroni-adjusted *p*=0·047). The difference in survival between the low and high PLS groups was more pronounced in males than in females.

#### Sex-specific Cox regression analyses

Overall, the PLS showed slightly more protective associations in males compared to females (Tables S1–S2). In Model 1, one standard deviation increase in the PLS was statistically significantly associated with a lower risk of all-cause mortality in both males (hazard ratio [HR]=0·800, 95% CI=0·745–0·860, difference in concordance index [ΔC-index]=0·009) and females (HR=0·887, 95% CI=0·814–0·968, ΔC-index=0·005). This association remained statistically significant across all models in both males (highest HR=0·836, 95% CI=0·777–0·899) and females (highest HR=0·909, 95% CI=0·834–0·990). The associations of lifestyle factors and education level were largely similar between the sexes. However, LTPA, former smoking, and education level were statistically significantly associated with all-cause mortality across all models in males but not in females. Additionally, heavy smoking had a stronger association with all-cause mortality in females (lowest HR=3·750, 95% CI=2·602–5·405; highest HR=4·075, 95% CI=2·890–5·746) than in males (lowest HR=3·281, 95% CI=2·588–4·161; highest HR=3·931, 95% CI=3·141–4·921). As individual predictors, alcohol consumption and smoking behaviour showed the largest contributions to the discriminatory ability for all-cause mortality in males, with ΔC-indices of 0·014 and 0·034, respectively (Table S5). In contrast, for females, BMI and smoking behaviour had the greatest discriminatory contributions (ΔC-indices: 0·007 and 0·034; Table S6).

The PLS did not have statistically significant interactions with lifestyle factors or education level in males (*p*>0·05). It did not have statistically significant interactions with LTPA; BMI; alcohol consumption; occasional, former, light, or heavy smoking; or education level (*p*>0·05), but it had a statistically significant interaction with medium smoking (highest *p*=0·011) in females. We suggest that this single interaction in a subset can most likely be explained by the uneven distribution of the participants in the smoking behaviour groups and did not conduct further time-dependent modelling.

Even though the proportional hazards assumption was statistically violated for alcohol consumption (in males) in some models, visual inspection of the Schoenfeld residuals did not indicate meaningful violations (Table S7). The proportional hazards assumption violations of the PCs were neglected, as the PCs were considered adjusting factors. No notable outliers were observed in the data based on the Martingale and deviance residuals. The linearity assumption was met. Statistical significance was set at a *p*-value of less than 0·05.

### Supplementary Tables

#### Table S1: Sex-specific descriptive statistics of the Finnish Twin Cohort study sample

|  | **Males** | | | **Females** | | |
| --- | --- | --- | --- | --- | --- | --- |
| **Variable** | **Mean (SD)** | **IQR** | **N** | **Mean (SD)** | **IQR** | **N** |
| **Age at entry (years)** | 57·82 (9·75) | 12·93 | 2522 | 57·13 (10·17) | 14·94 | 3054 |
| **Age at end of follow-up (years)** | 74·41 (7·80) | 12·03 | 2522 | 75·39 (7·92) | 12·82 | 3054 |
| **PLS (per 10^7^ units)** | -0·23 (0·96) | 1·32 | 2522 | -0·24 (0·95) | 1·29 | 3054 |
| **LTPA (MET-h/wk)** | 2·84 (2·62) | 2·47 | 2522 | 2·47 (1·82) | 1·92 | 3054 |
| **BMI (kg/m^2^)** | 25·59 (3·19) | 4·04 | 2521 | 24·51 (4·18) | 4·86 | 3052 |
| **Alcohol consumption (g/d)** | 14·60 (17·36) | 17·17 | 2522 | 5·39 (8·89) | 5·47 | 3051 |
| **Education level (years)** | 8·07 (3·03) | 4·00 | 2403 | 8·21 (2·92) | 4·00 | 3017 |
|  | **n** | **%** | **N** | **n** | **%** | **N** |
| **Smoking behaviour** |  |  | 2519 |  |  | 3054 |
| Never | 817 | 32·43 |  | 1851 | 60·61 |  |
| Occasional | 90 | 3·57 |  | 79 | 2·59 |  |
| Former | 870 | 34·54 |  | 517 | 16·93 |  |
| Light | 107 | 4·25 |  | 190 | 6·22 |  |
| Medium | 282 | 11·19 |  | 277 | 9·07 |  |
| Heavy | 353 | 14·01 |  | 140 | 4·58 |  |

The polygenic lifespan score (PLS) was z-standardized. SD=standard deviation. IQR=Interquartile range. n=number of participants in a category. N=number of participants. PLS=polygenic lifespan score. LTPA=leisure-time physical activity. MET=metabolic equivalent of task. BMI=body mass index.

#### Table S2: Associations of the polygenic lifespan score and lifestyle factors with the risk of all-cause mortality in males in the Older Finnish Twin Cohort

| **Events/Participants** | **Model 1**  807/2522 | **Model 2**  807/2522 | **Model 3**  807/2521 | **Model 4**  807/2522 | **Model 5**  806/2519 | **Model 6**  806/2518 | **Model 7**  773/2403 | **Model 8**  772/2399 |
| --- | --- | --- | --- | --- | --- | --- | --- | --- |
| **PLS** | **0·800**  **(0·745–0·860)** | **0·806**  **(0·750–0·865)** | **0·803**  **(0·748–0·863)** | **0·799**  **(0·742–0·859)** | **0·825**  **(0·767–0·886)** | **0·826**  **(0·769–0·888)** | **0·814**  **(0·757–0·875)** | **0·836**  **(0·777–0·899)** |
| **LTPA (MET-h/week)** |  | **0·936**  **(0·905–968)** |  |  |  | **0·952**  **(0·920–0·985)** |  | **0·957**  **(0·924–0·990)** |
| **BMI (kg/m^2^)** |  |  | **1·036**  **(1·013–1·060)** |  |  | **1·034**  **(1·011–1·059)** |  | **1·030**  **(1·006–1·055)** |
| **Alcohol consumption (g/d)** |  |  |  | **1·016**  **(1·013–1·020)** |  | **1·011**  **(1·008–1·015)** |  | **1·011**  **(1·007–1·014)** |
| **Smoking behaviour** |  |  |  |  |  |  |  |  |
| Never |  |  |  |  | 1·000 (ref). | 1·000 (ref). |  | 1·000 (ref). |
| Occasional |  |  |  |  | 1·291  (0·831–2·004) | 1·161  (0·746–1·807) |  | 1·241  (0·792–1·946) |
| Former |  |  |  |  | **1·464**  **(1·210–1·772)** | **1·282**  **(1·055–1·558)** |  | **1·291**  **(1·059–1·573)** |
| Light |  |  |  |  | **2·145**  **(1·467–3·137)** | **1·998**  **(1·364–2·928)** |  | **1·887**  **(1·278–2·787)** |
| Medium |  |  |  |  | **2·306**  **(1·791–2·970)** | **2·116**  **(1·639–2·731)** |  | **2·130**  **(1·647–2·755)** |
| Heavy |  |  |  |  | **3·931**  **(3·141–4·921)** | **3·224**  **(2·549–4·076)** |  | **3·281**  **(2·588–4·161)** |
| **Education level** |  |  |  |  |  |  | **0·956**  **(0·929–0·983)** | **0·965**  **(0·938–0·993)** |

PLS=polygenic lifespan score. LTPA=leisure-time physical activity. MET=metabolic equivalent of task. BMI=body mass index. The table includes the hazard ratios and their 95% confidence intervals for each covariate inspected in the different models. Model 1=all-cause mortality ~ PLS+ the first ten genetic principal components of ancestry + family relatedness as a random factor. Model 2=Model 1 + LTPA. Model 3=Model 1 + BMI. Model 4=Model 1 + alcohol consumption. Model 5=Model 1 + smoking behaviour. Model 6 =Model 1 + lifestyle covariates. Model 7=Model 1 + education level. Model 8=Model 1 + all covariates. Bold type indicates statistical significance at the level of *p*<0·05.

#### Table S3: Associations of the polygenic lifespan score and lifestyle factors with the risk of all-cause mortality in females in the Older Finnish Twin Cohort

| **Events/Participants** | **Model 1**  598/3054 | **Model 2**  598/3054 | **Model 3**  597/3052 | **Model 4**  597/3051 | **Model 5**  598/3054 | **Model 6**  596/5567 | **Model 7**  595/3017 | **Model 8**  593/3012 |
| --- | --- | --- | --- | --- | --- | --- | --- | --- |
| **PLS** | **0·887**  **(0·814–0·968)** | **0·886**  **(0·813–0·967)** | **0·897**  **(0·822–0·978)** | **0·888**  **(0·815–0·968)** | **0·899**  **(0·825–0·980)** | **0·908**  **(0·834–0·989)** | **0·889**  **(0·815–0·970)** | **0·909**  **(0·834–0·990)** |
| **LTPA (MET-h/week)** |  | 0·986  (0·939–1·036) |  |  |  | 1·001  (0·954–1·051) |  | 0·999  (0·951–1·049) |
| **BMI (kg/m^2^)** |  |  | **1·034**  **(1·014–1·055)** |  |  | **1·038**  **(1·017–1·058)** |  | **1·036**  **(1·015–1·057)** |
| **Alcohol consumption (g/d)** |  |  |  | **1·018**  **(1·010–1·026)** |  | 1·008  (0·999–1·016) |  | **1·009**  **(1·000–1·017)** |
| **Smoking behaviour** |  |  |  |  |  |  |  |  |
| Never |  |  |  |  | 1·00 (ref.) | 1·00 (ref.) |  | 1·000 (ref.) |
| Occasional |  |  |  |  | 1·406  (0·824–2·397) | 1·353  (0·794–2·307) |  | 1·406  (0·824–2·401) |
| Former |  |  |  |  | 1·214  (0·937–1·574) | 1·205  (0·928–1·565) |  | 1·228  (0·944–1·596) |
| Light |  |  |  |  | 1·432  (0·971–2·112) | 1·456  (0·986–2·149) |  | **1·479**  **(1·001–2·184)** |
| Medium |  |  |  |  | **2·061**  **(1·528–2·780)** | **2·060**  **(1·521–2·791)** |  | **2·036**  **(1·502–2·762)** |
| Heavy |  |  |  |  | **4·075**  **(2·890–5·746)** | **3·747**  **(2·604–5·391)** |  | **3·750**  **(2·602–5·405)** |
| **Education level** |  |  |  |  |  |  | 0·969  (0·935–1·003) | 0·977  (0·943–1·012) |

PLS=polygenic lifespan score. LTPA=leisure-time physical activity. MET=metabolic equivalent of task. BMI=body mass index. The table includes the hazard ratios and their 95% confidence intervals for each covariate inspected in the different models. Model 1=all-cause mortality ~ PLS+ the first ten genetic principal components of ancestry + family relatedness as a random factor. Model 2=Model 1 + LTPA. Model 3=Model 1 + BMI. Model 4=Model 1 + alcohol consumption. Model 5=Model 1 + smoking behaviour. Model 6 =Model 1 + lifestyle covariates. Model 7=Model 1 + education level. Model 8=Model 1 + all covariates. Bold type indicates statistical significance at the level of *p*<0·05.

#### Table S4: Bivariate linear mixed-effects associations of the PLS and other fixed-effects covariates

| **Variables associated with the polygenic lifespan score** | **Beta coefficient** | **Standard error** | ***p*** |
| --- | --- | --- | --- |
| **Sex** | -0·007 | 0·032 | 0·825 |
| **Leisure-time physical activity (MET-h/wk)** | 0·012 | 0·012 | 0·301 |
| **Body mass index (kg/m^2^)** | -0·053 | 0·012 | **<0·05** |
| **Alcohol consumption (g/d)** | -0·006 | 0·012 | 0·632 |
| **Smoking behaviour** | - | - | - |
| Never | 0·000 (ref.) | 0·000 (ref.) | 0·000 (ref.) |
| Occasional | 0·067 | 0·064 | 0·300 |
| Former | 0·018 | 0·027 | 0·507 |
| Light | -0·004 | 0·050 | 0·942 |
| Medium | -0·108 | 0·038 | **0·004** |
| Heavy | -0·096 | 0·040 | **0·016** |
| **Education level** | 0·085 | 0·014 | **<0·050** |
| **Principal component 1** | 0·073 | 0·016 | **<0·050** |
| **Principal component 2** | -0·018 | 0·014 | 0·216 |
| **Principal component 3** | -0·012 | 0·015 | 0·421 |
| **Principal component 4** | 0·008 | 0·014 | 0·561 |
| **Principal component 5** | 0·021 | 0·015 | 0·178 |
| **Principal component 6** | -0·011 | 0·015 | 0·456 |
| **Principal component 7** | -0·008 | 0·014 | 0·549 |
| **Principal component 8** | 0·035 | 0·015 | **0·021** |
| **Principal component 9** | -0·008 | 0·015 | 0·614 |
| **Principal component 10** | -0·018 | 0·015 | 0·246 |

MET=metabolic equivalent of task. The results are from bivariate linear mixed-effects models, where all continuous variables were standardised. The results for the smoking behaviour groups were from the associations between the PLS and binary smoking behaviour group variables. Family relatedness (family number) was a random intercept. The number of participants in each model was 5411. Statistical significance at the level of *p*<0·05 is signified using bold type.

#### Table S5: Concordance index differences between the full model and models excluding study variables separately for males

|  | **C-index without the variable (SE)** | **ΔC-index** |
| --- | --- | --- |
| **PLS** | 0·658 (0·011) | 0·009 |
| **LTPA** | 0·664 (0·011) | 0·003 |
| **BMI** | 0·666 (0·011) | 0·001 |
| **Alcohol consumption** | 0·653 (0·011) | 0·014 |
| **Smoking behaviour** | 0·633 (0·011) | 0·034 |
| Never | ref. | ref. |
| Occasional | 0·666 (0·011) | 0·000 |
| Former | 0·665 (0·011) | 0·001 |
| Light | 0·662 (0·011) | 0·004 |
| Medium | 0·655 (0·011) | 0·012 |
| Heavy | 0·640 (0·011) | 0·026 |
| **Education level** | 0·666 (0·011) | 0·001 |

C-index=concordance index. SE=standard error. ΔC-index=difference in C-index between the full model and a model without a covariate. PLS=polygenic lifespan score. LTPA=leisure-time physical activity. BMI=body mass index. The C-index for the model (full model), including all covariates and the first ten genetic principal components of ancestry, was 0·666 (SE=0·011). The number of events was 772 and the number of participants was 2399.

#### Table S6: Concordance index differences between the full model and models excluding study variables separately for females

|  | **C-index without the variable (SE)** | **ΔC-index** |
| --- | --- | --- |
| **PLS** | 0·603 (0·014) | 0·005 |
| **LTPA** | 0·608 (0·014) | 0·000 |
| **BMI** | 0·601 (0·013) | 0·007 |
| **Alcohol consumption** | 0·605 (0·014) | 0·003 |
| **Smoking behaviour** | 0·574 (0·014) | 0·034 |
| Never | ref. | ref. |
| Occasional | 0·606 (0·014) | 0·002 |
| Former | 0·606 (0·014) | 0·002 |
| Light | 0·603 (0·014) | 0·005 |
| Medium | 0·590 (0·014) | 0·018 |
| Heavy | 0·590 (0·013) | 0·018 |
| **Education level** | 0·607 (0·014) | 0·001 |

C-index=concordance index. SE=standard error. ΔC-index=difference in C-index between the full model and a model without a covariate. PLS=polygenic lifespan score. LTPA=leisure-time physical activity. BMI=body mass index. The C-index for the model (full model), including all covariates and the first ten genetic principal components of ancestry, was 0·608 (SE=0·014). The number of events was 593 and the number of participants was 3012.

#### Table S7: Compendium of the proportional hazards assumption test results of all variables for all participants, males, and females.

|  | **All** | | |  | **Males** | | |  | | **Female** | | |
| --- | --- | --- | --- | --- | --- | --- | --- | --- | --- | --- | --- | --- |
|  | **χ^2^ (min–max)** | **df** | ***p* (min–max)** |  | ***χ* ^2^ (min–max)** | **df** | ***p* (min–max)** |  | ***χ* ^2^ (min–max)** | | **df** | ***p* (min–max)** |
| **PLS** | 0·0085–0·53 | 1 | 0·47–0·93 |  | 0·15–0·20 | 1 | 0·65–0·90 |  | 0·37–0·75 | | 1 | 0·39–0·54 |
| **Sex** | 0·068–0·81 | 1 | 0·37–0·79 |  | ·· | ·· | ·· |  | ·· | | ·· | ·· |
| **LTPA** | 0·086–0·52 | 1 | 0·47–0·77 |  | 0·0035–0·23 | 1 | 0·63–0·95 |  | 0·32–0·54 | | 1 | 0·46–0·57 |
| **BMI** | 0·027–0·063 | 1 | 0·80–0·87 |  | 0·42–1·66 | 1 | 0·20–0·52 |  | 0·26–0·44 | | 1 | 0·51–0·61 |
| **Alcohol consumption** | 0·60–1·29 | 1 | 0·26–0·44 |  | 2·91–4·11 | 1 | **0·043**–0·088 |  | 0·023–0·049 | | 1 | 0·82–0·88 |
| **Smoking behaviour** | 7·96–9·26 | 5 | 0·099–0·16 |  | 3·60–4·83 | 5 | 0·44–0·61 |  | 5·86–5·94 | | 5 | 0·31–0·32 |
| Occasional | 0·22–0·59 | 1 | 0·44–0·64 |  | 0·50–1·71 | 1 | 0·19–0·48 |  | 0·00055–0·40 | | 1 | 0·53–0·98 |
| Former | 0·095–0·36 | 1 | 0·55–0·76 |  | 0·34–0·73 | 1 | 0·39–0·56 |  | 0·36–0·45 | | 1 | 0·50–0·55 |
| Light | 3·46–4·59 | 1 | **0·032**–0·062 |  | 0·69–2·31 | 1 | 0·13–0·41 |  | 2·70–3·14 | | 1 | 0·076–0·10 |
| Medium | 3·80–5·04 | 1 | **0·025**–0·51 |  | 0·13–2·42 | 1 | 0·12–0·72 |  | 0·000017–2·78 | | 1 | 0·095–1·00 |
| Heavy | 0·00039–0·083 | 1 | 0·77–0·98 |  | 0·10–0·24 | 1 | 0·62–0·75 |  | 0·00034–0·069 | | 1 | 0·79–0·99 |
| **Education level** | 0·029–0·32 | 1 | 0·57–0·87 |  | 0·00044–0·16 | 1 | 0·69–0·98 |  | 0·0098–0·069 | | 1 | 0·56–0·79 |
| **PC1** | 0·048–3·34 | 1 | 0·068–0·83 |  | 0·0016–0·12 | 1 | 0·73–0·97 |  | 6·55–7·82 | | 1 | **0·0052–0·011** |
| **PC2** | 2·38–3·55 | 1 | 0·060–0·12 |  | 0·019–0·22 | 1 | 0·64–0·89 |  | 3·14–4·10 | | 1 | **0·043–**0·076 |
| **PC3** | 2·40–4·24 | 1 | **0·040**–0·12 |  | 2·35–4·05 | 1 | **0·044**–0·13 |  | 0·79–1·07 | | 1 | 0·30–0·37 |
| **PC4** | 0·30–2·38 | 1 | 0·12–0·59 |  | 0·43–2·08 | 1 | 0·15–0·51 |  | 0·00044–0·026 | | 1 | 0·87–0·98 |
| **PC5** | 0·54–1·56 | 1 | 0·21–0·46 |  | 6·64–9·29 | 1 | **0·0023–0·010** |  | 0·53–1·12 | | 1 | 0·29–0·47 |
| **PC6** | 1·16–2·91 | 1 | 0·088–0·28 |  | 3·07–6·70 | 1 | **0·0096–0·080** |  | 0·00032–0·10 | | 1 | 0·75–0·99 |
| **PC7** | 0·000080–3·39 | 1 | 0·066–0·99 |  | 0·37–1·13 | 1 | 0·29–0·85 |  | 0·081–0·26 | | 1 | 0·61–0·78 |
| **PC8** | 0·0086–0·87 | 1 | 0·35–0·93 |  | 0·45–1·53 | 1 | 0·22–0·50 |  | 0·00086–0·051 | | 1 | 0·82–0·98 |
| **PC9** | 0·53–0·92 | 1 | 0·34–0·47 |  | 0·88–1·72 | 1 | 0·19–0·35 |  | 0·073–0·13 | | 1 | 0·72–0·79 |
| **PC10** | 0·029–0·53 | 1 | 0·47–0·87 |  | 0·41–1·02 | 1 | 0·31–0·52 |  | 0·65–1·01 | | 1 | 0·31–0·42 |
| **Global** | 0·14–27·20 | 21 | 0·062–15·36 |  | 17·92–31·17 | 20 | **0·022**–0·12 |  | 14·01–23·09 | | 20 | 0·13–0·30 |

***χ*** ^2^=chi-square. Df=degrees of freedom. PLS=polygenic lifespan score. LTPA=leisure-time physical activity. BMI=body mass index. The ***χ*** ^2^- and *p*-values of the proportional hazards assumption test results are the minimum and maximum values among all computed Cox regression models. Bold type indicates statistical significance at the level of *p*<0·05.

#### Table S8: Descriptive statistics of the complete case analyses in the Older Finnish Twin Cohort

|  | **All (N=5411)** | |  | **Males (N=2399)** | |  | **Females (N*=*3012)** | |
| --- | --- | --- | --- | --- | --- | --- | --- | --- |
|  | **Mean (SD)** | **IQR** |  | **Mean (SD)** | **IQR** |  | **Mean (SD)** | **IQR** |
| **Age at follow-up entry (years)** | 57·41 (9·99) | 13·99 |  | 57·77 (9·77) | 1·31 |  | 57·13 (10·16) | 14·95 |
| **Age at follow-up end (years)** | 74·95 (7·88) | 12·50 |  | 74·38 (7·80) | 12·95 |  | 75·40 (7·92) | 12·83 |
| **PLS (per 10^7^ units)** | -0·23 (0·95) | 1·30 |  | -0·23 (0·96) | 12·07 |  | -0·23 (0·95) | 1·29 |
| **LTPA (MET-h/wk)** | 2·63 (2·22) | 2·17 |  | 2·83 (2·63) | 2·45 |  | 2·47 (1·82) | 1·92 |
| **BMI (kg/m^2^)** | 24·99 (3·81) | 4·62 |  | 25·59 (3·19) | 4·07 |  | 24·51 (4·17) | 4·87 |
| **Alcohol consumption (g/d)** | 9·45 (14·11) | 9·03 |  | 14·56 (17·43) | 17·05 |  | 5·38 (8·87) | 5·47 |
| **Education level (years)** | 8·15 (2·97) | 4·00 |  | 8·07 (3·03) | 4·00 |  | 8·21 (2·92) | 4·00 |
|  | **n** | **%** |  | **n** | **%** |  | **n** | **%** |
| **Smoking behaviour** | ·· | ·· |  | ·· | ·· |  | ·· | ·· |
| Never | 2600 | 48·05 |  | 775 | 32·31 |  | 1825 | 60·59 |
| Occasional | 161 | 2·98 |  | 83 | 3·46 |  | 78 | 2·59 |
| Former | 1336 | 24·69 |  | 826 | 34·43 |  | 510 | 16·93 |
| Light | 287 | 5·30 |  | 100 | 4·17 |  | 187 | 6·21 |
| Medium | 548 | 10·13 |  | 273 | 11·38 |  | 275 | 9·13 |
| Heavy | 479 | 8·85 |  | 342 | 14·26 |  | 137 | 4·55 |

The polygenic lifespan score (PLS) was z-standardized. SD=standard deviation. IQR=Interquartile range. n=number of participants in a category. N=number of participants. PLS=polygenic lifespan score. LTPA=leisure-time physical activity. MET=metabolic equivalent of task. BMI=body mass index.

#### Table S9. Complete case analysis of the associations of the polygenic lifespan score and lifestyle factors with the risk of all-cause mortality in all participants in the Older Finnish Twin Cohort

| **Events/Participants** | **Model 1**  1365/5411 | **Model 2**  1365/5411 | **Model 3**  1365/5411 | **Model 4**  1365/5411 | **Model 5**  1365/5411 | **Model 6**  1365/5411 | **Model 7**  1365/5411 | **Model 8**  1365/5411 | **Model 9**  1365/5411 |
| --- | --- | --- | --- | --- | --- | --- | --- | --- | --- |
| **PLS** | **0 841**  **(0 794–0 891)** | **0 837**  **(0 792–0 885)** | **0 841**  **(0 795–0 891)** | **0 848**  **(0 801–0 898)** | **0 839**  **(0 792–0 889)** | **0 864**  **(0 817–0 913)** | **0 865**  **(0 818–0 915)** | **0 844**  **(0 797–0 894)** | **0 863**  **(0 816–0 912)** |
| **Sex** | ·· | **0 498**  **(0 443–0 559)** | ·· | ·· | ·· | ·· | ·· | ·· | **0 678**  **(0 597–0 770)** |
| **LTPA (MET-h/week)** | ·· | ·· | **0 958**  **(0 931–0 986)** | ·· | ·· | ·· | **0 969**  **(0 942–0 997)** | ·· | **0 968**  **(0 941–0 995)** |
| **BMI (kg/m^2^)** | ·· | ·· | ·· | **1 039**  **(1 024–1 055)** | ·· | ·· | **1 037**  **(1 021–1 052)** | ·· | **1 031**  **(1 015–1 047)** |
| **Alcohol consumption (g/d)** | ·· | ·· | ·· | ·· | **1 021**  **(1 018–1 024)** | ·· | **1 013**  **(1 010–1 016)** | ·· | **1 011**  **(1 007–1 014)** |
| **Smoking behaviour** | ·· | ·· | ·· | ·· | ·· | ·· | ·· | ·· | ·· |
| Never | ·· | ·· | ·· | ·· | ·· | 1·000 (ref.) | 1·000 (ref.) | ·· | 1·000 (ref.) |
| Occasional | ·· | ·· | ·· | ·· | ·· | **1 510**  **(1 072–2 128)** | **1 369**  **(0 971–1 929)** | ·· | **1 278**  **(0 907–1 801)** |
| Former | ·· | ·· | ·· | ·· | ·· | **1 656**  **(1 434–1 913)** | **1 456**  **(1 257–1 687)** | ·· | **1 277**  **(1 097–1 488)** |
| Light | ·· | ·· | ·· | ·· | ·· | **1 786**  **(1 357–2 350)** | **1 737**  **(1 320–2 286)** | ·· | **1 666**  **(1 267–2 189)** |
| Medium | ·· | ·· | ·· | ·· | ·· | **2 473**  **(2 044–2 993)** | **2 303**  **(1 900–2 791)** | ·· | **2 078**  **(1 713–2 522)** |
| Heavy | ·· | ·· | ·· | ·· | ·· | **4 872**  **(4 072–5 829)** | **3 851**  **(3 181–4 663)** | ·· | **3 341**  **(2 751–4 056)** |
| **Education level** | ·· | ·· | ·· | ·· | ·· | ·· | ·· | **0 959**  **(0 797–0 894)** | **0 970**  **(0 948–0 991)** |

PLS=polygenic lifespan score. LTPA=leisure-time physical activity. MET=metabolic equivalent of task. BMI=body mass index. The table includes the hazard ratios and their 95% confidence intervals for each covariate inspected in the different models. Model 1=all-cause mortality ~ PLS + the first ten genetic principal components of ancestry + family relatedness as random factor. Model 2=Model 1 + sex. Model 3=Model 1 + LTPA. Model 4=Model 1 + BMI. Model 5=Model 1 + alcohol consumption. Model 6=Model 1 + smoking behaviour. Model 7=Model 1 + lifestyle covariates. Model 8=Model 1 + education level. Model 9=Model 1 + all covariates. Bold type indicates statistical significance at the level of *p*<0·05.

#### Table S10. Complete case analysis of the associations of the polygenic lifespan score and lifestyle factors with the risk of all-cause mortality in males in the Older Finnish Twin Cohort

| **Events/Participants** | **Model 1**  772/2399 | **Model 2**  772/2399 | **Model 3**  772/2399 | **Model 4**  772/2399 | **Model 5**  772/2399 | **Model 6**  772/2399 | **Model 7**  772/2399 | **Model 8**  772/2399 |
| --- | --- | --- | --- | --- | --- | --- | --- | --- |
| **PLS** | **0·805**  **(0·748–0·865)** | **0·810**  **(0·754–0·870)** | **0·808**  **(0·752–0·868)** | **0·802**  **(0·744–0·864)** | **0·833**  **(0·775–0·896)** | **0·834**  **(0·776–0·897)** | **0·809**  **(0·753–0·870)** | **0·836**  **(0·778–0·900)** |
| **LTPA (MET-h/week)** | ·· | **0·936**  **(0·905–0·969)** | ·· | ·· | ·· | **0·953**  **(0·920–0·986)** | ·· | **0·957**  **(0·924–0·990)** |
| **BMI (kg/m^2^)** | ·· | ·· | **1·035**  **(1·011–1·060)** | ·· | ·· | **1·033**  **(1·009–1·058)** | ·· | **1·030**  **(1·006–1·055)** |
| **Alcohol consumption (g/d)** | ·· | ·· | ·· | **1·016**  **(1·013–1·019)** | ·· | **1·011**  **(1·007–1·014)** | ·· | **1·011**  **(1·007–1·014)** |
| **Smoking behaviour** | ·· | ·· | ·· | ·· | ·· | ·· | ·· | ·· |
| Never | ·· | ·· | ·· | ·· | 1·000 (ref.) | 1·000 (ref.) | ·· | 1·000 (ref.) |
| Occasional | ·· | ·· | ·· | ·· | 1343  (0·859–2·102) | 1·209  (0·771–1·896) | ·· | 1·241  (0·792–1·946) |
| Former | ·· | ·· | ·· | ·· | **1·475**  **(1·215–1·792)** | **1·297**  **(1·064–1·582)** | ·· | **1·291**  **(1·059–1·573)** |
| Light | ·· | ·· | ·· | ·· | **2·065**  **(1·400–3·046)** | **1·923**  **(1·301–2·842)** | ·· | **1·887**  **(1·278–2·787)** |
| Medium | ·· | ·· | ·· | ·· | **2·366**  **(1·834–3·053)** | **2·173**  **(1·680–2·810)** | ·· | **2·130**  **(1·647–2·755)** |
| Heavy | ·· | ·· | ·· | ·· | **3·998**  **(3·185–5·020)** | **3·306**  **(2·607–4·192)** | ·· | **3·281**  **(2·588–4·161)** |
| **Education level** | ·· | ·· | ·· | ·· | ·· | ·· | **0·955**  **(0**·**929–0·983)** | **0·965**  **(0·938–0·993)** |

PLS=polygenic lifespan score. LTPA=leisure-time physical activity. MET=metabolic equivalent of task. BMI=body mass index. The table includes the hazard ratios and their 95% confidence intervals for each covariate inspected in the different models. Model 1=all-cause mortality ~ PLS+ the first ten genetic principal components of ancestry + family relatedness as a random factor. Model 2=Model 1 + LTPA. Model 3=Model 1 + BMI. Model 4=Model 1 + alcohol consumption. Model 5=Model 1 + smoking behaviour. Model 6=Model 1 + lifestyle covariates. Model 7=Model 1 + education level. Model 8=Model 1 + all covariates. Bold type indicates statistical significance at the level of *p*<0·05.

#### Table S11: Complete case analysis of the associations of the polygenic lifespan score and lifestyle factors with the risk of all-cause mortality in females in the Older Finnish Twin Cohort

| **Events/Participants** | **Model 1**  593/3012 | **Model 2**  593/3012 | **Model 3**  593/3012 | **Model 4**  593/3012 | **Model 5**  593/3012 | **Model 6**  593/3012 | **Model 7**  593/3012 | **Model 8**  593/3012 |
| --- | --- | --- | --- | --- | --- | --- | --- | --- |
| **PLS** | **0·889**  **(0·815–0·970)** | **0·888**  **(0·814–0·969)** | **0·898**  **(0·823–0·980)** | **0·889**  **(0·815–0·969)** | **0·901**  **(0·827–0·982)** | **0·909**  **(0·834–0·990)** | **0·890**  **(0·816–0·971)** | **0·909**  **(0·834–0·990)** |
| **LTPA (MET-h/week)** | ·· | 0·985  (0·938–1·035) | ·· | ·· | ·· | 0·998  (0·950–1·048) | ·· | 0·999  (0·951–1·049) |
| **BMI (kg/m^2^)** | ·· | ·· | **1·035**  **(1·014–1·056)** | ·· | ·· | **1·038**  **(1·17–1·058)** | ·· | **1·036**  **(1·015–1·057)** |
| **Alcohol consumption (g/d)** | ·· | ·· | ·· | **1·029**  **(1·010–1·026)** | ·· | 1·008  (0·999–1·017) | ·· | **1·009**  **(1·000–1·017)** |
| **Smoking behaviour** | ·· | ·· | ·· | ·· | ·· | ·· | ·· | ·· |
| Never | ·· | ·· | ·· | ·· | 1·000 (ref.) | 1·000 (ref.) | ·· | 1·000 (ref.) |
| Occasional | ·· | ·· | ·· | ·· | 1·439  (0·842–2·459) | 1·397  (0·819–2·385) | ·· | 1·406  (0·824–2·401) |
| Former | ·· | ·· | ·· | ·· | 1·230  (0·948–1·596) | 1·215  (0·935–1·580) | ·· | 1·228  (0·944–1·596) |
| Light | ·· | ·· | ·· | ·· | **1·460**  **(0·989–2·156)** | **1·486**  **(1·006–2·196)** | ·· | **1·479**  **(1·001–2·184)** |
| Medium | ·· | ·· | ·· | ·· | **2·044**  **(1·511–2·765)** | **2·061**  **(1·52–2·794)** | ·· | **2·036**  **(1·502–2·762)** |
| Heavy | ·· | ·· | ·· | ·· | **4·149**  **(2·937–5·860)** | **3·795**  **(2·633–5·468)** | ·· | **3·750**  **(2·602–5·405)** |
| **Education level** | ·· | ·· | ·· | ·· | ·· | ·· | **0·968**  **(0·935–1·003)** | **0·977**  **(0·943–1·012)** |

PLS=polygenic lifespan score. LTPA=leisure-time physical activity. MET=metabolic equivalent of task. BMI=body mass index. The table includes the hazard ratios and their 95% confidence intervals for each covariate inspected in the different models. Model 1=all-cause mortality ~ PLS+ the first ten genetic principal components of ancestry + family relatedness as a random factor. Model 2=Model 1 + LTPA. Model 3=Model 1 + BMI. Model 4=Model 1 + alcohol consumption. Model 5=Model 1 + smoking behaviour. Model 6=Model 1 + lifestyle covariates. Model 7=Model 1 + education level. Model 8=Model 1 + all covariates. Bold type indicates statistical significance at the level of *p*<0·05.

#### Table S12: Associations of sex, the lifestyle factors, and education level with the risk of all-cause mortality in females in the Older Finnish Twin Cohort

| **Deaths/Participants** | **Model 1**  1405/5576 | | **Model 2**  1405/5576 | | **Model 3**  1405/5573 | | **Model 4**  1404/5573 | | **Model 5**  1404/5573 | | **Model 6**  1402/5567 | | **Model 7**  1368/5420 | | **Model 8**  1365/5411 | |
| --- | --- | --- | --- | --- | --- | --- | --- | --- | --- | --- | --- | --- | --- | --- | --- | --- |
| **Sex** | | **0·498**  **(0·444–0·560)** | | ·· | | ·· | | ·· | | ·· | | ·· | | ·· | | **0·678**  **(0·597–0·770)** |
| **LTPA (MET-h/week)** | | ·· | | **0·959**  **(0·932–0·986)** | | ·· | | ·· | | ·· | | **0·970**  **(0·943–0·998)** | | ·· | | **0·968**  **(0·941–0·995)** |
| **BMI (kg/m^2^)** | | ·· | | ·· | | **1·042**  **(1·027–1·057)** | | ·· | | ·· | | **1·039**  **(1·024–1·054)** | | ·· | | **1·032**  **(1·016–1·048)** |
| **Alcohol consumption (g/d)** | | ·· | | ·· | | ·· | | **1·021**  **(1·018–1·024)** | | ·· | | **1·013**  **(1·010–1·016)** | | ·· | | **1·010**  **(1·007–1·014)** |
| **Smoking behaviour** | | ·· | | ·· | | ·· | | ·· | | ·· | | ·· | | ·· | | ·· |
| Never | | ·· | | ·· | | ·· | | ·· | | 1·000 (ref.) | | 1·000 (ref.) | | ·· | | 1·000 (ref.) |
| Occasional | | ·· | | ·· | | ·· | | ·· | | **1·437**  **(1·023–2·016)** | | 1·294  (0·920–1·818) | | ·· | | 1·265  (0·896–1·786) |
| Former | | ·· | | ·· | | ·· | | ·· | | **1·667**  **(1·446–1·922)** | | **1·465**  **(1·266–1·695)** | | ·· | | **1·297**  **(1·113–1·512)** |
| Light | | ·· | | ·· | | ·· | | ·· | | **1·827**  **(1·393–2·397)** | | **1·780**  **(1·356–2·336)** | | ·· | | **1·693**  **(1·286–2·229)** |
| Medium | | ·· | | ·· | | ·· | | ·· | | **2·537**  **(2·100–3·066)** | | **2·361**  **(1·950–2·859)** | | ·· | | **2·166**  **(1·784–2·629)** |
| Heavy | | ·· | | ·· | | ·· | | ·· | | **4·858**  **(4·068–5·802)** | | **3·821**  **(3·160–4·620)** | | ·· | | **3·392**  **(2·792–4·122)** |
| **Education level** | | ·· | | ·· | | ·· | | ·· | | ·· | | ·· | | **0·955**  **(0·934–0·977)** | | **0·967**  **(0·946–0·988)** |

LTPA=leisure-time physical activity. BMI=body mass index. The table includes the hazard ratios and their 95% confidence intervals for each covariate inspected in the different models. All models include family relatedness a random factor. The first ten genetic principal components were not considered. Model 1=all-cause mortality ~ sex. Model 2= all-cause mortality ~ LTPA. Model 3 = all-cause mortality ~ BMI. Model 4=all-cause mortality ~ alcohol consumption. Model 5=all-cause mortality ~ smoking behaviour. Model 6= all-cause mortality ~ all lifestyle covariates. Model 7=all-cause mortality ~ education level. Model 8= all-cause mortality ~ all covariates. Bold type indicates statistical significance at the level of *p*<0·05.

### **Supplementary Figures**

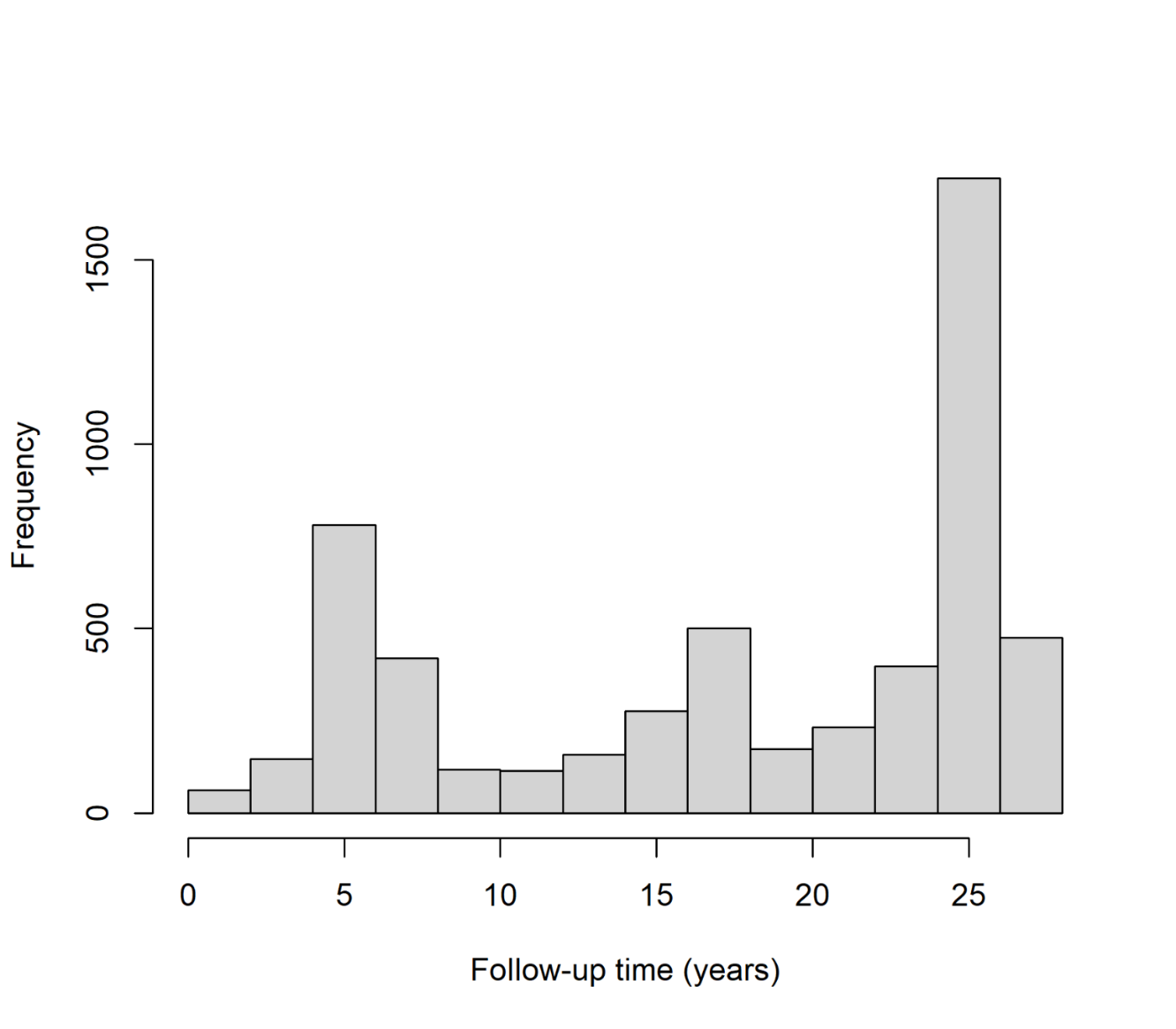

#### **Figure S1: Distribution of follow-up times in the study.**

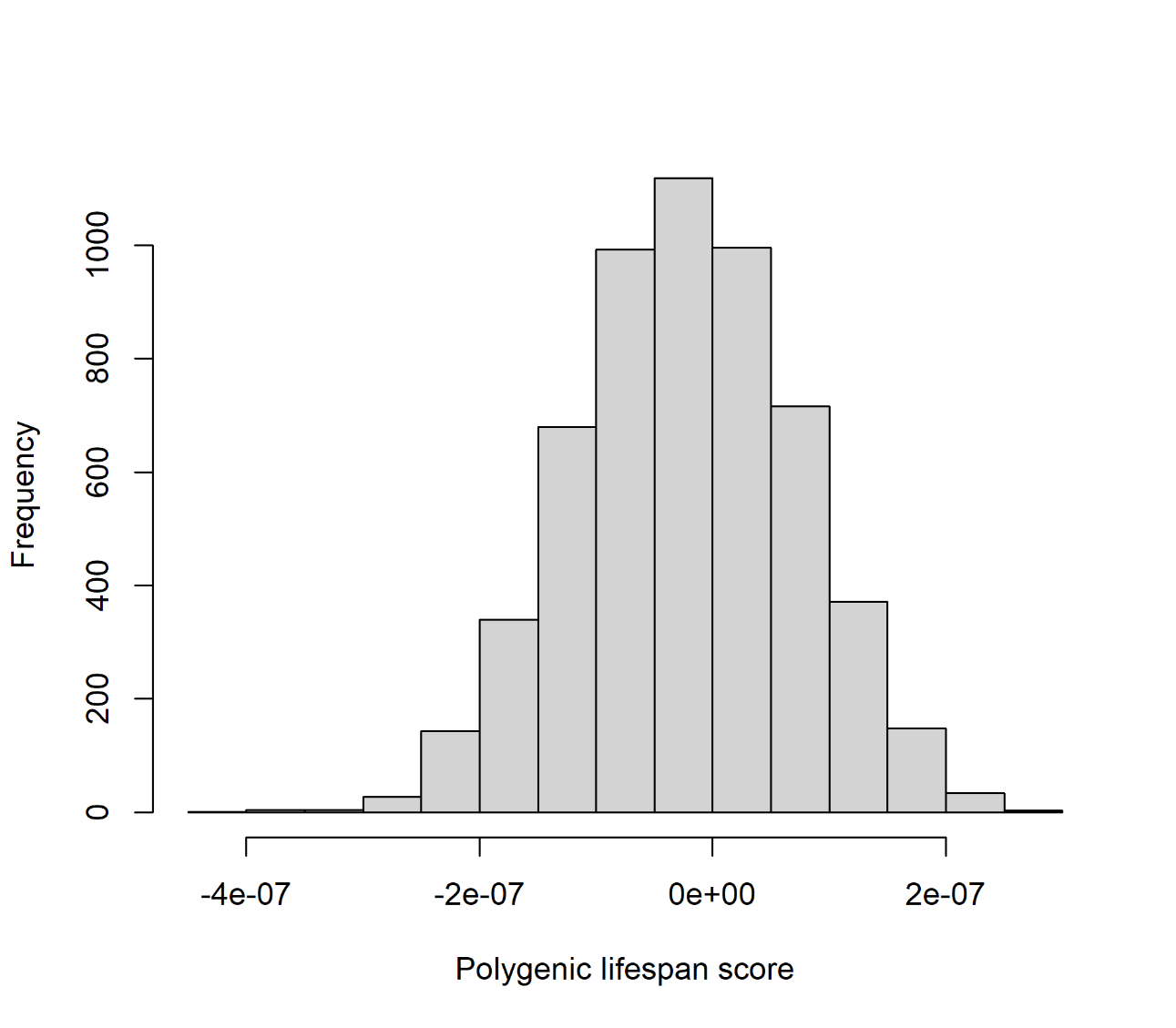

#### Figure S2: Distribution of the polygenic lifespan score.

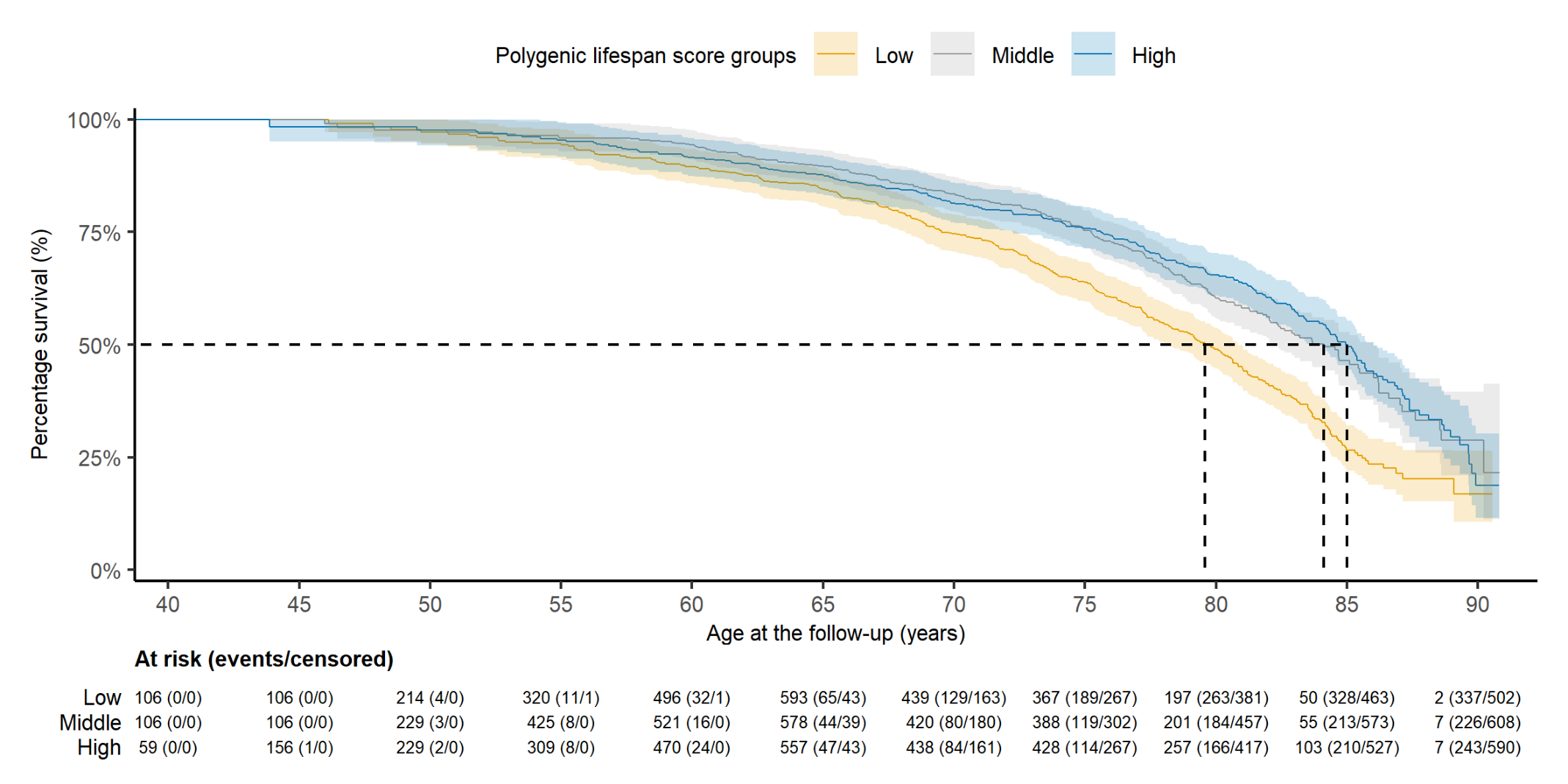

Figure S3: Kaplan–Meier survival curves for all-cause mortality risk in relation to the polygenic lifespan score groups for males in the Older Finnish Twin Cohort.

Median survival is indicated using a dashed line.

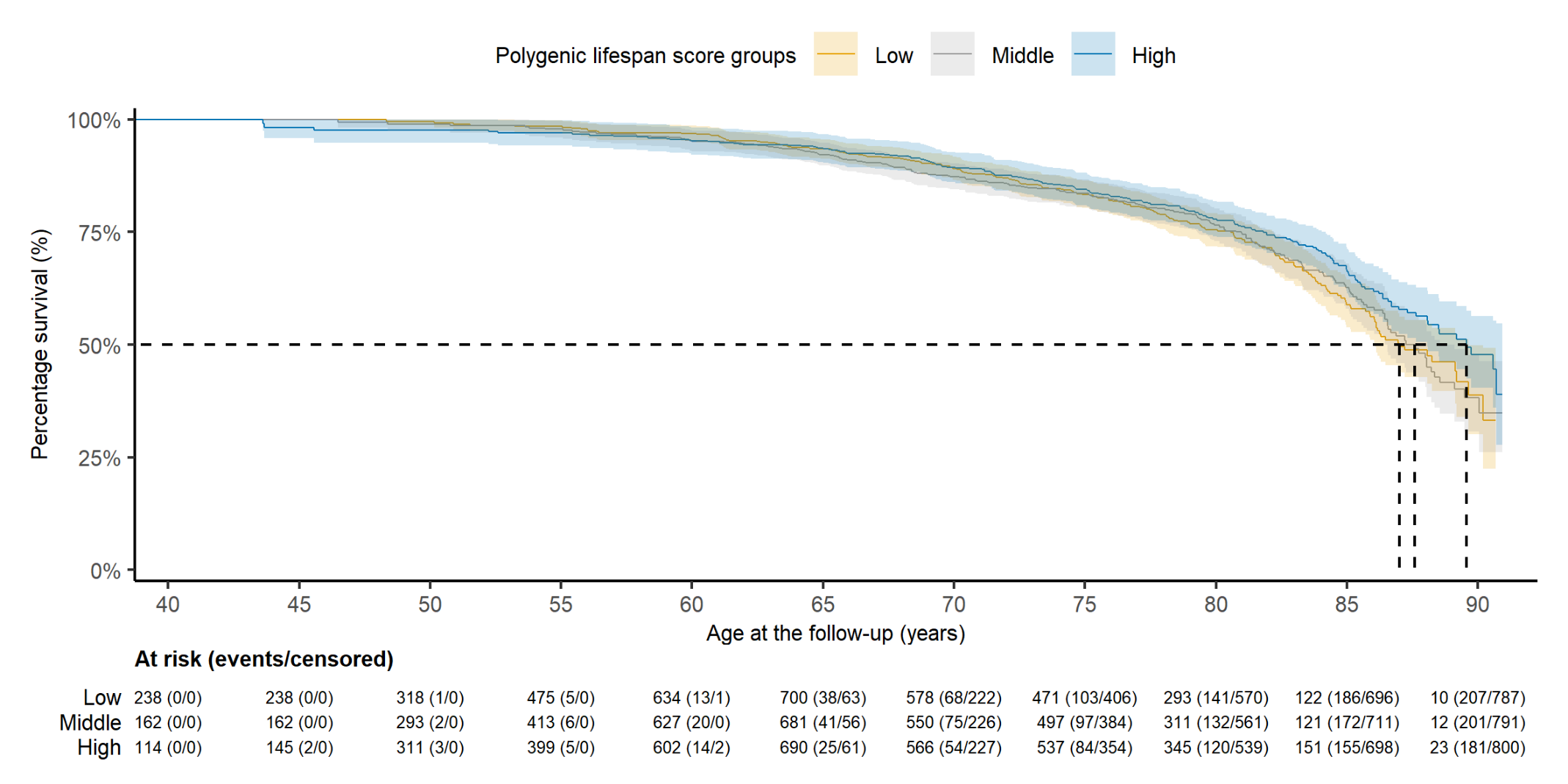

Figure S4: Kaplan–Meier survival curves for all-cause mortality risk in relation to the polygenic lifespan score groups for females in the Older Finnish Twin Cohort.

Median survival is indicated using a dashed line.

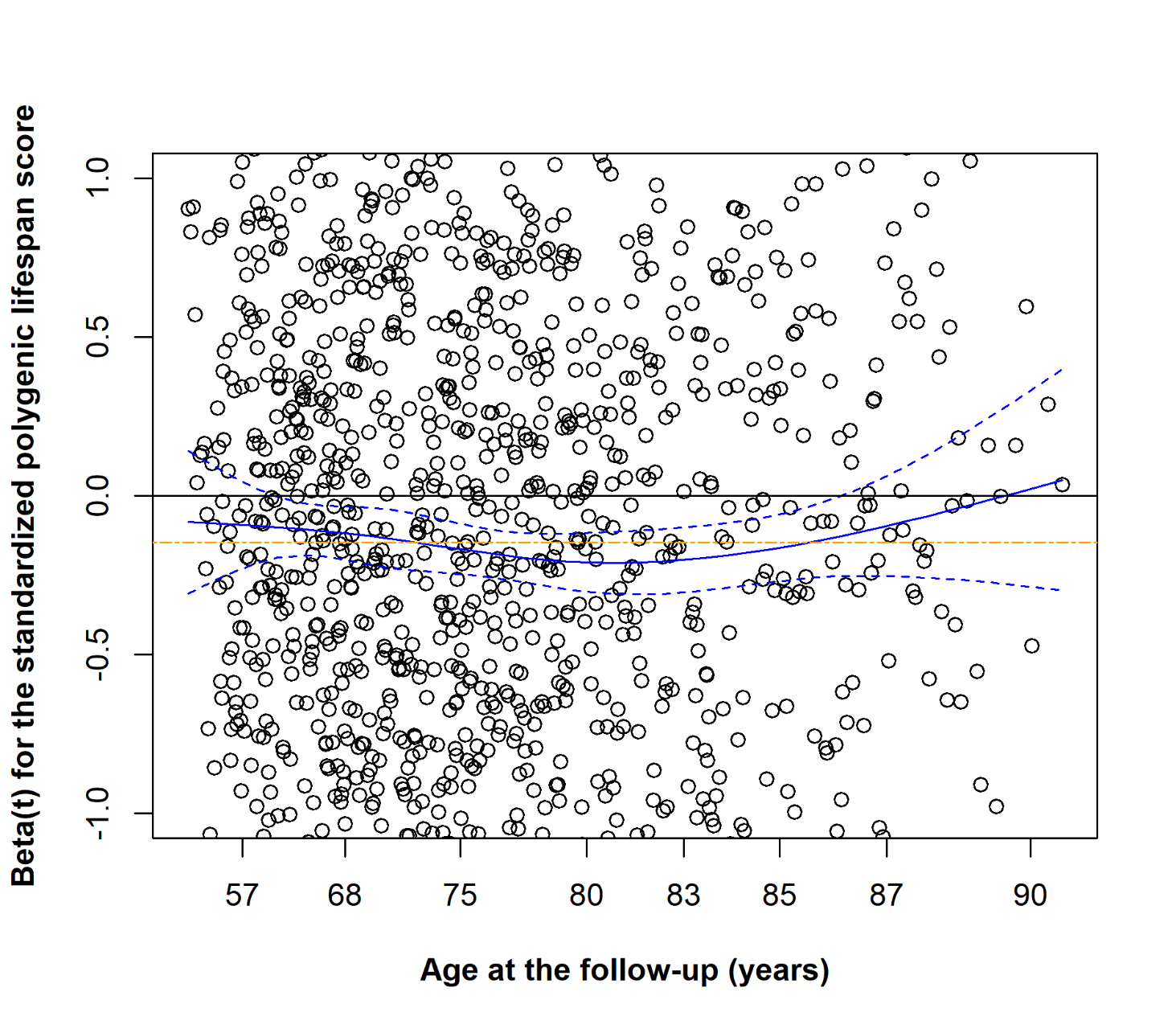

Figure S5: Schoenfeld residual plot of the association between the standardised polygenic lifespan score and the risk of all-cause mortality in the full model.

The association of the polygenic lifespan score with all-cause mortality was adjusted for the first ten genetic principal components of ancestry and family relatedness. The model also included sex, leisure-time physical activity, body mass index, alcohol consumption, smoking behaviour, and education level as covariates. The solid black line is the reference line for zero residual deviation. The solid blue line indicates a smoothing spline of the fit and the dashed blue lines indicate a ±2-standard-error band around the fit. The two-dash orange line represents the *β*_1_ reference line for the polygenic lifespan score in the model. Each circle represents a scaled Schoenfeld residual for an individual event (observation), illustrating the deviation from the proportional hazards assumption over time.

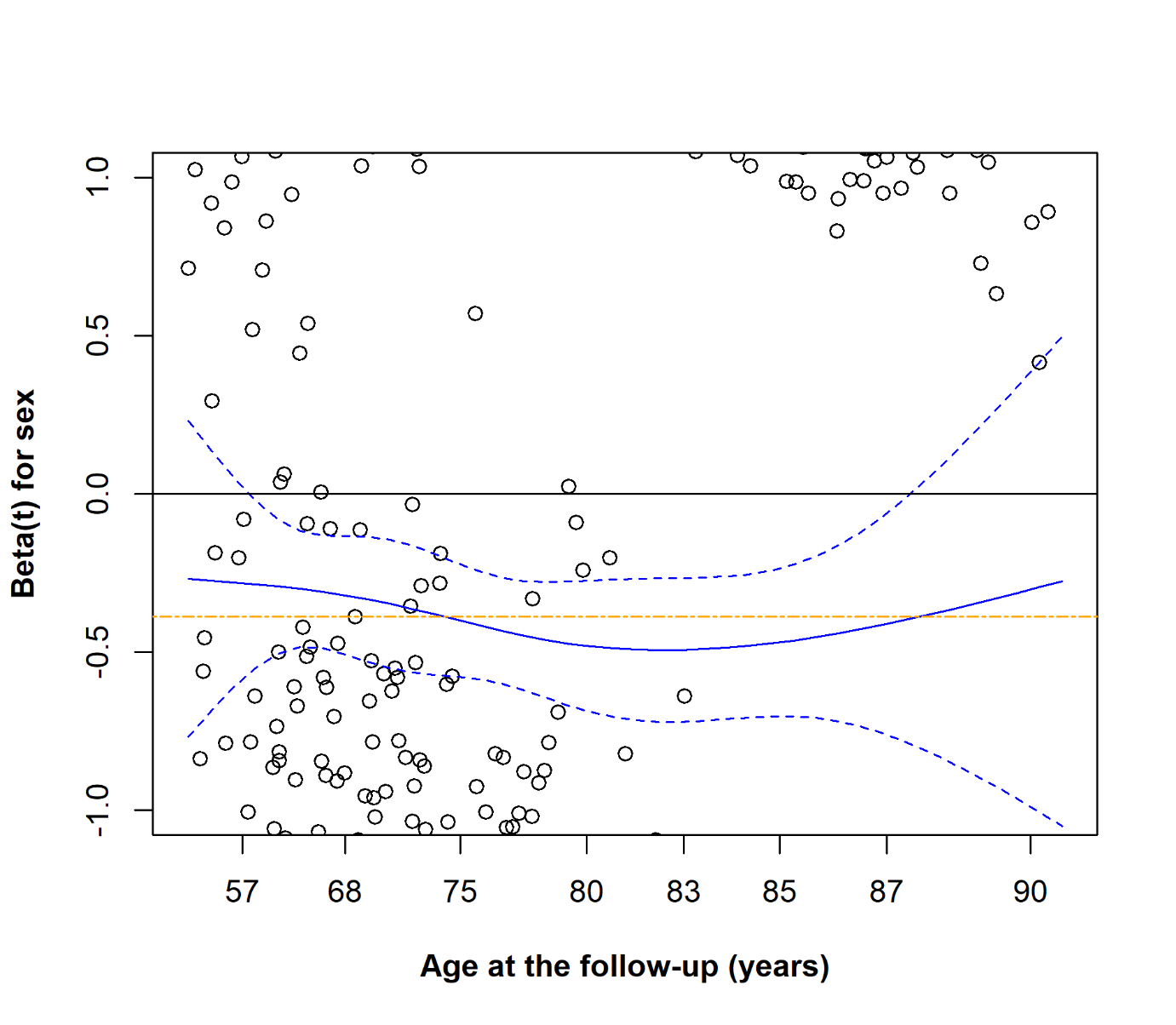

Figure S6: Schoenfeld residual plot of the association between sex and the risk of all-cause mortality in the full model.

The association of sex with all-cause mortality was adjusted for the first ten genetic principal components of ancestry and family relatedness. The model also included the polygenic lifespan score, leisure-time physical activity, body mass index, alcohol consumption, smoking behaviour, and education level as covariates. The solid black line is the reference line for zero residual deviation. The solid blue line indicates a smoothing spline of the fit and the dashed blue lines indicate a ±2-standard-error band around the fit. The two-dash orange line represents the *β*_2_ reference line of sex in the model. Each circle represents a scaled Schoenfeld residual for an individual event (observation), illustrating the deviation from the proportional hazards assumption over time.

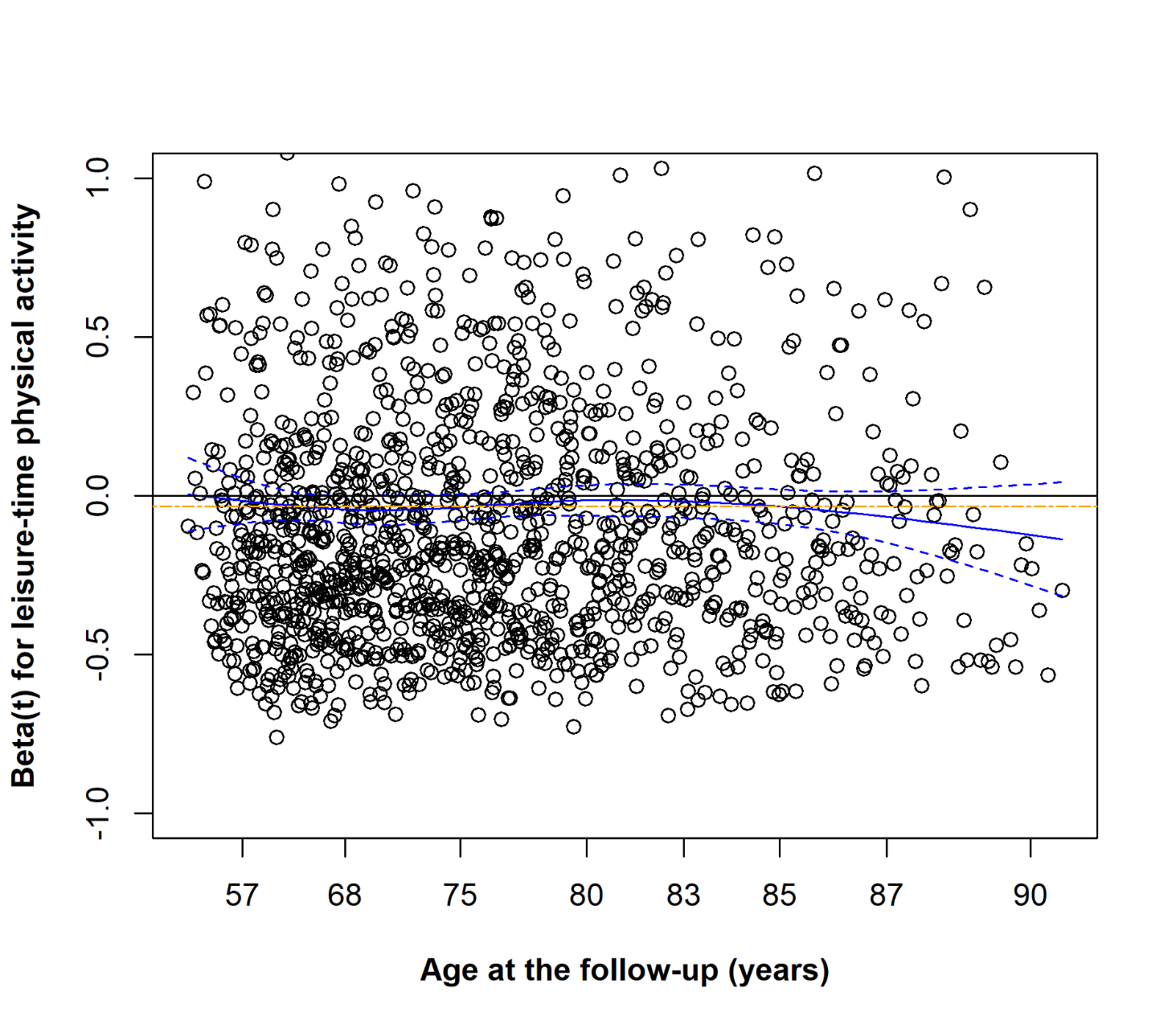

Figure S7: Schoenfeld residual plot of the association between leisure-time physical activity and the risk of all-cause mortality in the full model.

The association of leisure-time physical activity with all-cause mortality was adjusted for the first ten genetic principal components of ancestry and family relatedness. The model also included the polygenic lifespan score, body mass index, alcohol consumption, smoking behaviour, and education level as covariates. The solid black line is the reference line for zero residual deviation. The solid blue line indicates a smoothing spline of the fit and the dashed blue lines indicate a ±2-standard-error band around the fit. The two-dash orange line represents the *β*_3_ reference line of leisure-time physical activity in the model. Each circle represents a scaled Schoenfeld residual for an individual event (observation), illustrating the deviation from the proportional hazards assumption over time.

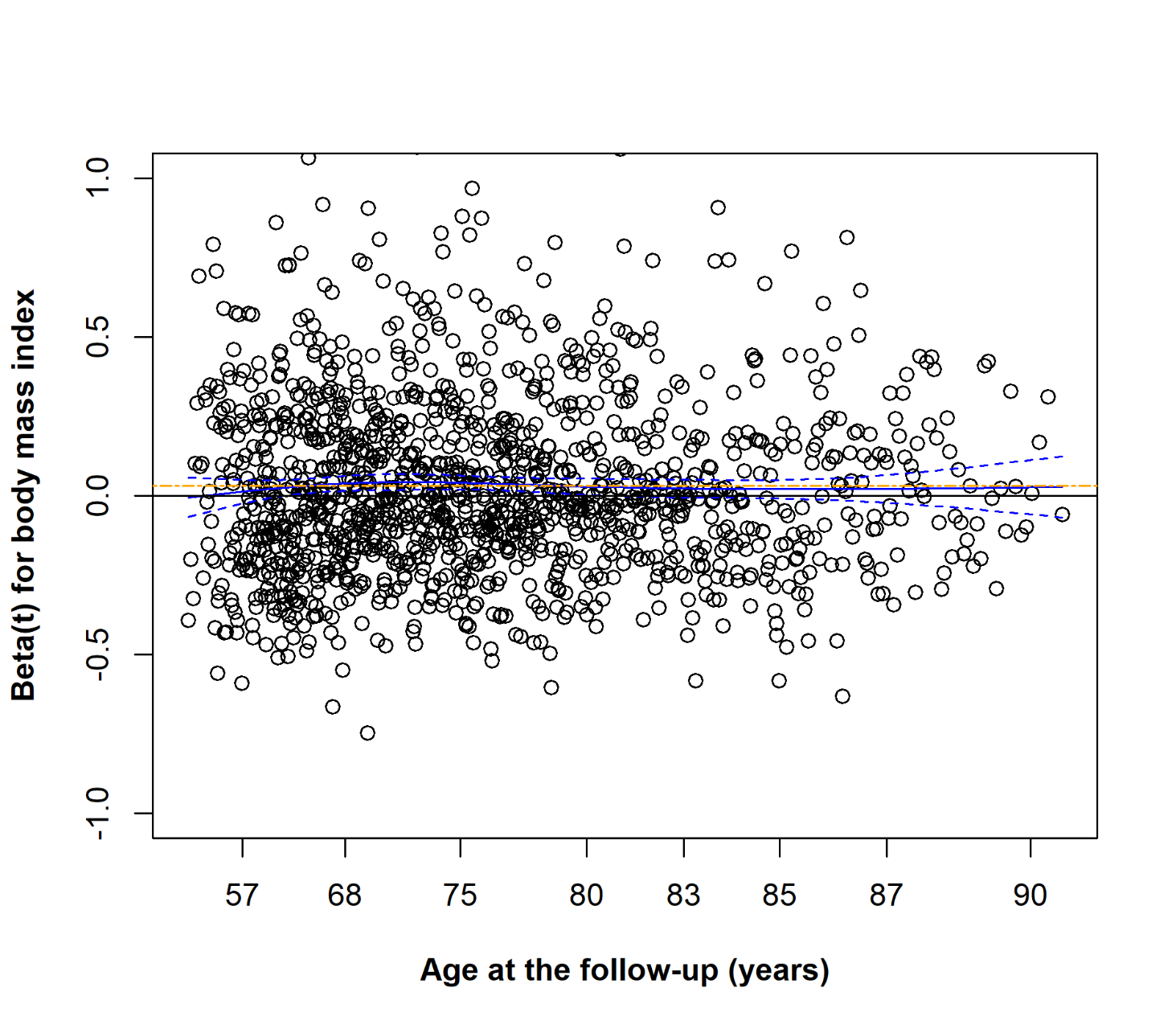

Figure S8: Schoenfeld residual plot of the association between body mass index and the risk of all-cause mortality in the full model.

The association of body mass index with all-cause mortality was adjusted for the first ten genetic principal components of ancestry and family relatedness. The model also included the polygenic lifespan score, leisure-time physical activity, alcohol consumption, smoking behaviour, and education level as covariates. The solid black line is the reference line for zero residual deviation. The solid blue line indicates a smoothing spline of the fit and the dashed blue lines indicate a ±2-standard-error band around the fit. The two-dash orange line represents the *β*_4_ reference line of body mass index in the model. Each circle represents a scaled Schoenfeld residual for an individual event (observation), illustrating the deviation from the proportional hazards assumption over time.

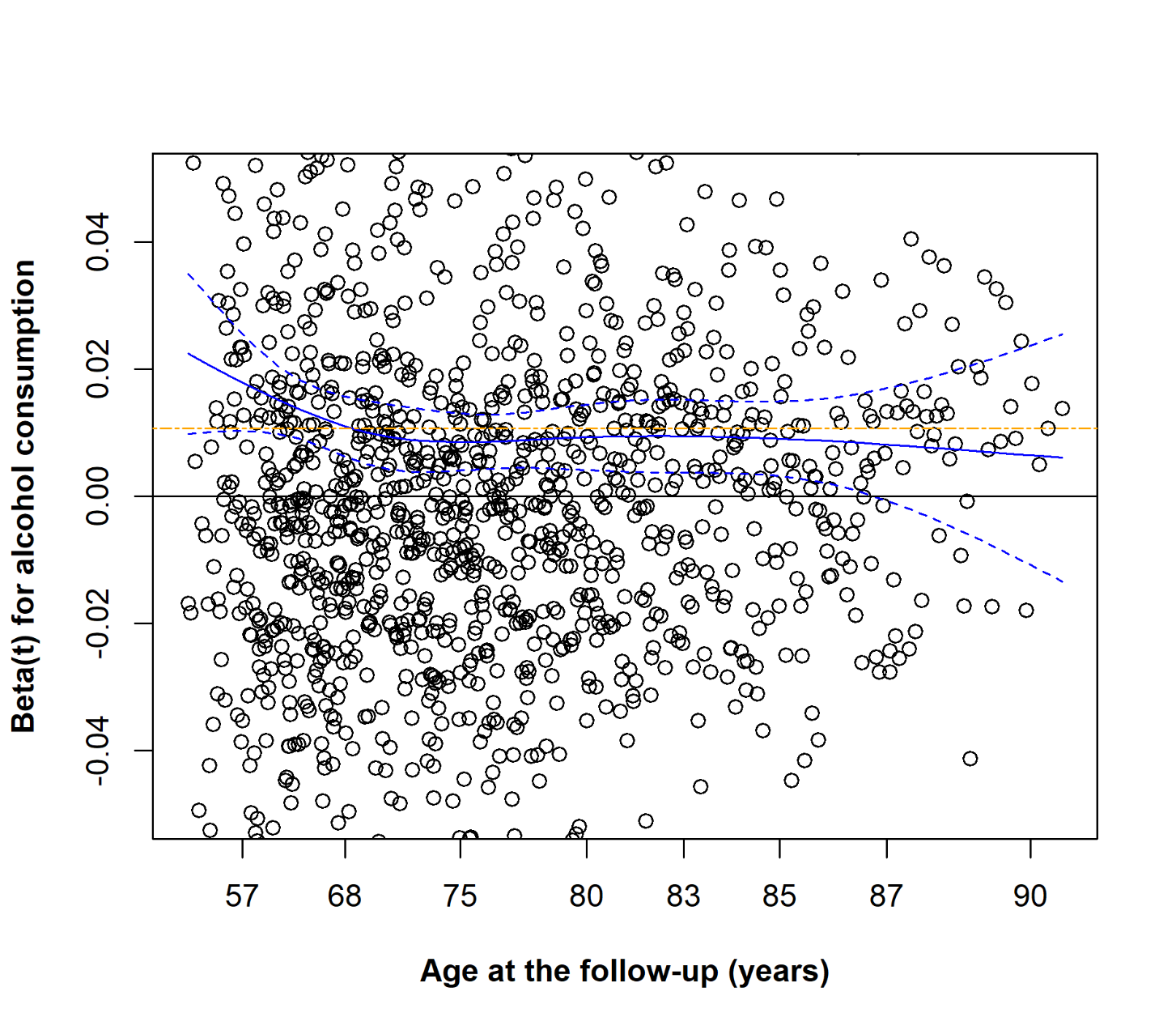

Figure S9: Schoenfeld residual plot of the association between alcohol consumption and the risk of all-cause mortality in the full model.

The association of alcohol consumption with all-cause mortality was adjusted for the first ten genetic principal components of ancestry and family relatedness. The model also included the polygenic lifespan score, leisure-time physical activity, body mass index, smoking behaviour, and education level as covariates. The solid black line is the reference line for zero residual deviation. The solid blue line indicates a smoothing spline of the fit and the dashed blue lines indicate a ±2-standard-error band around the fit. The two-dash orange line represents the *β*_5_ reference line of alcohol consumption in the model. Each circle represents a scaled Schoenfeld residual for an individual event (observation), illustrating the deviation from the proportional hazards assumption over time.

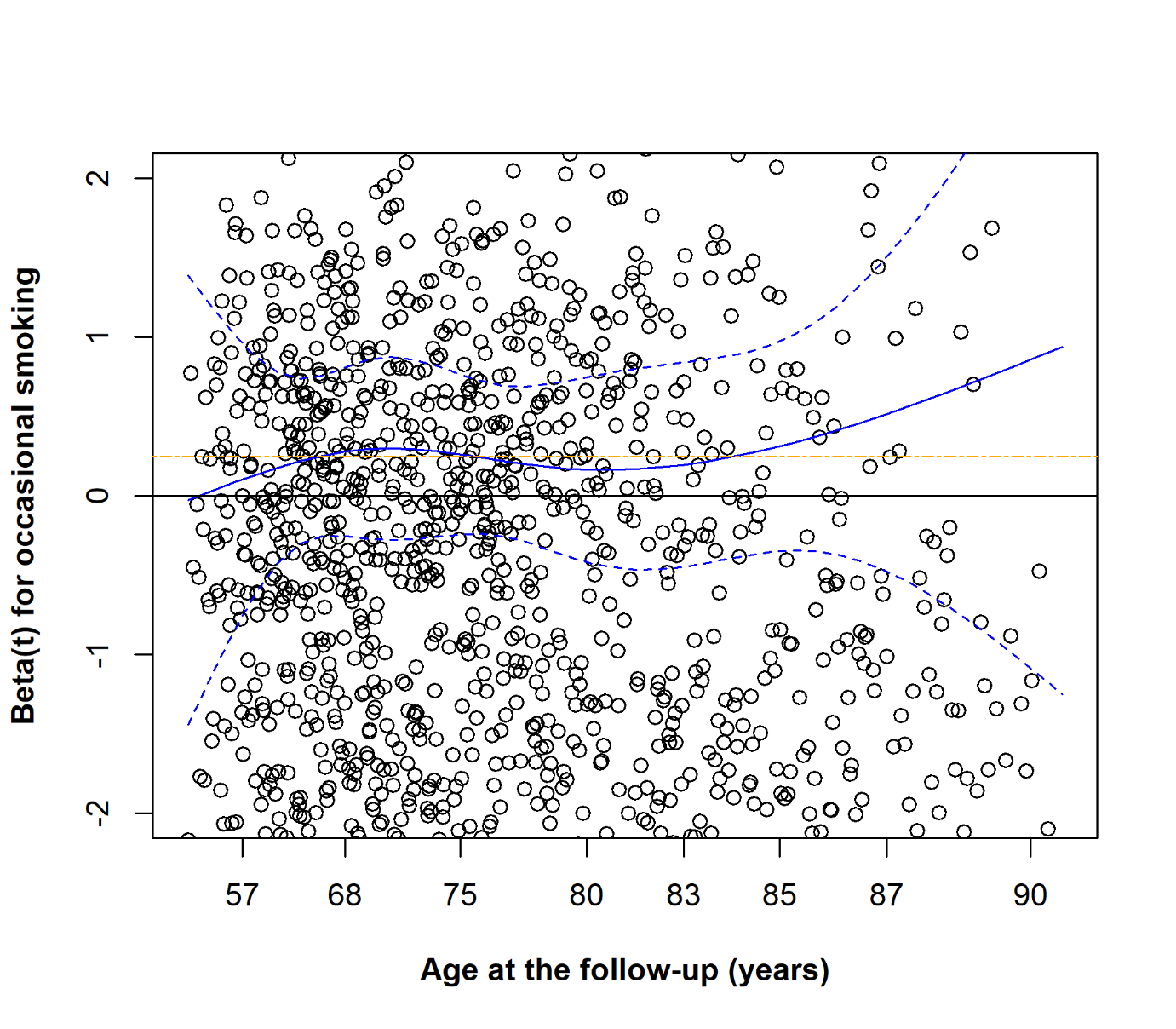

Figure S10: Schoenfeld residual plot of the association between the occasional smoking group and the risk of all-cause mortality in the full model.

The association of the occasional smoking group with all-cause mortality was adjusted for the first ten genetic principal components of ancestry and family relatedness. The model also included the polygenic lifespan score, leisure-time physical activity, body mass index, alcohol consumption, former smoking group, light smoking group, medium smoking group, heavy smoking group, and education level as covariates. The Schoenfeld residuals for each smoking behaviour group were obtained from models in which each group was recoded as a binary covariate (group vs. never smoking group). The solid black line is the reference line for zero residual deviation. The solid blue line indicates a smoothing spline of the fit and the dashed blue lines indicate a ±2-standard-error band around the fit. The two-dash orange line represents the *β_6_* reference line of heavy smoking in the model. Each circle represents a scaled Schoenfeld residual for an individual event (observation), illustrating the deviation from the proportional hazards assumption over time.

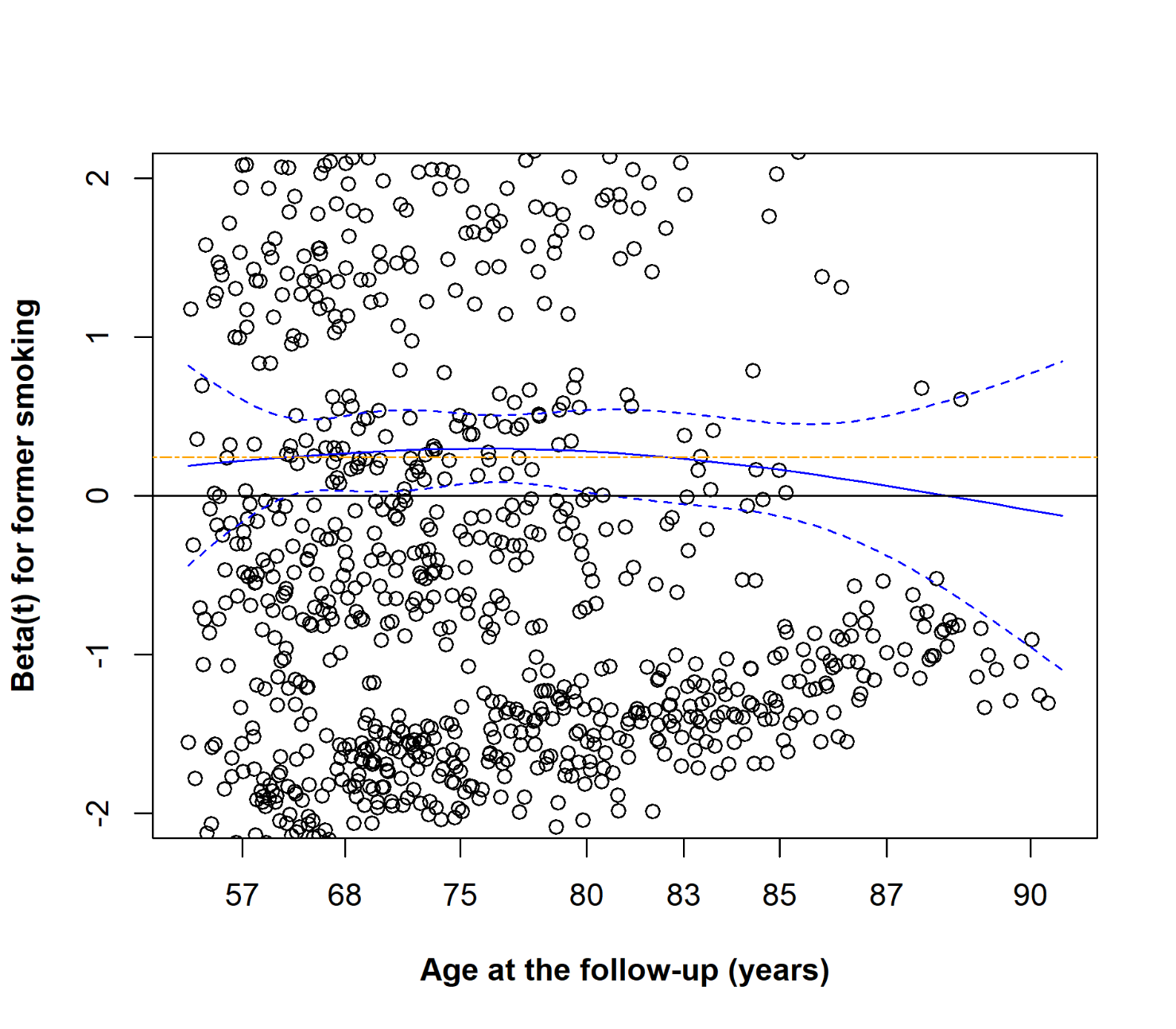

Figure S11: Schoenfeld residual plot of the association between the former smoking group and the risk of all-cause mortality in the full model.

The association of the former smoking group with all-cause mortality was adjusted for the first ten genetic principal components of ancestry and family relatedness. The model also included the polygenic lifespan score, leisure-time physical activity, body mass index, alcohol consumption, occasional smoking group, light smoking group, medium smoking group, heavy smoking group, and education level as covariates. The Schoenfeld residuals for each smoking behaviour group were obtained from models in which each group was recoded as a binary covariate (group vs. never smoking group). The solid black line is the reference line for zero residual deviation. The solid blue line indicates a smoothing spline of the fit and the dashed blue lines indicate a ±2-standard-error band around the fit. The two-dash orange line represents the *β_7_* reference line of heavy smoking in the model. Each circle represents a scaled Schoenfeld residual for an individual event (observation), illustrating the deviation from the proportional hazards assumption over time.

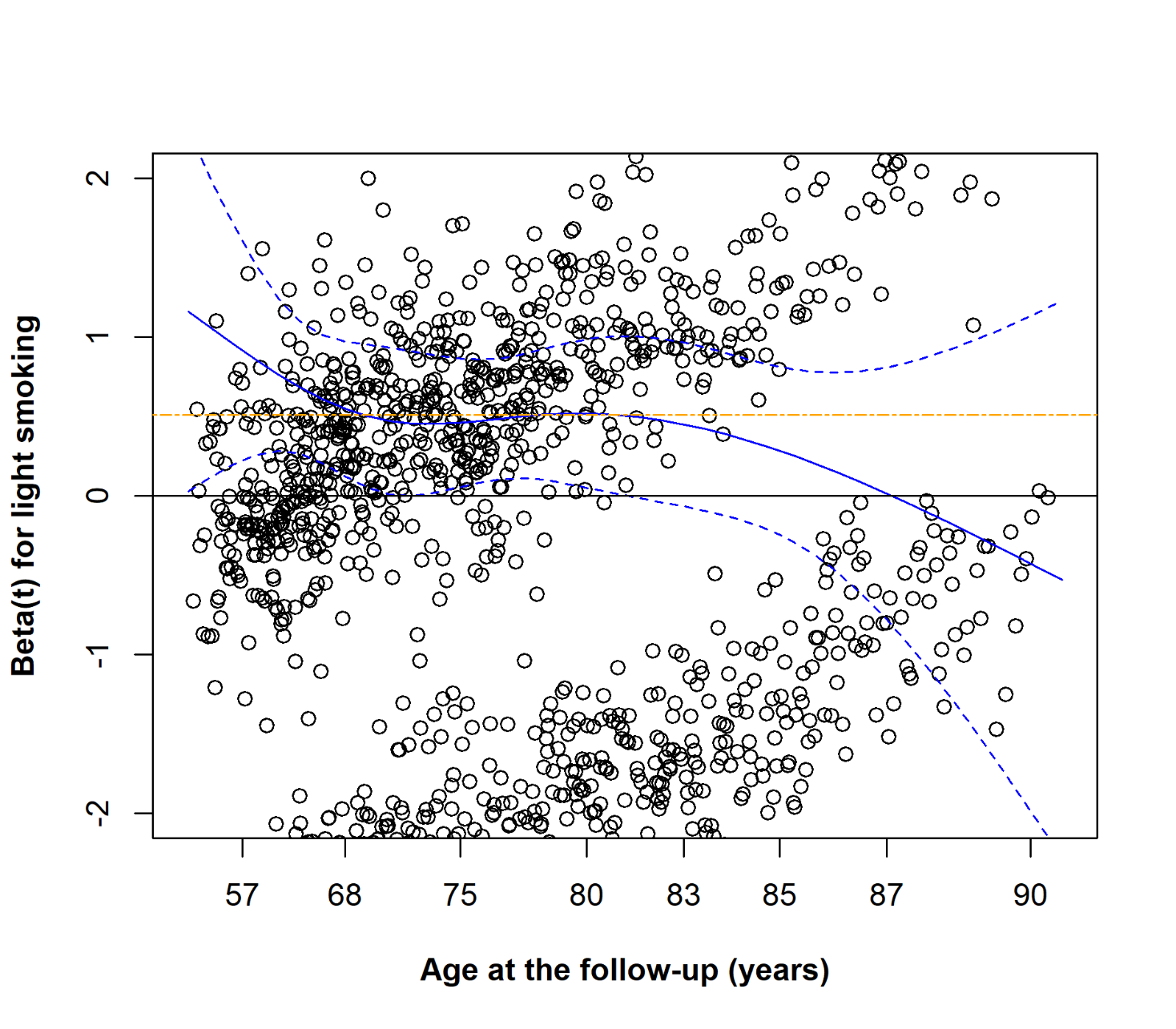

Figure S12: Schoenfeld residual plot of the association between the light smoking group and the risk of all-cause mortality in the full model.

The association of the former smoking group with all-cause mortality was adjusted for the first ten genetic principal components of ancestry and family relatedness. The model also included the polygenic lifespan score, leisure-time physical activity, body mass index, alcohol consumption, occasional smoking group, former smoking group, medium smoking group, heavy smoking group, and education level as covariates. The Schoenfeld residuals for each smoking behaviour group were obtained from models in which each group was recoded as a binary covariate (group vs. never smoking group). The solid black line is the reference line for zero residual deviation. The solid blue line indicates a smoothing spline of the fit and the dashed blue lines indicate a ±2-standard-error band around the fit. The two-dash orange line represents the *β_8_* reference line of heavy smoking in the model. Each circle represents a scaled Schoenfeld residual for an individual event (observation), illustrating the deviation from the proportional hazards assumption over time.

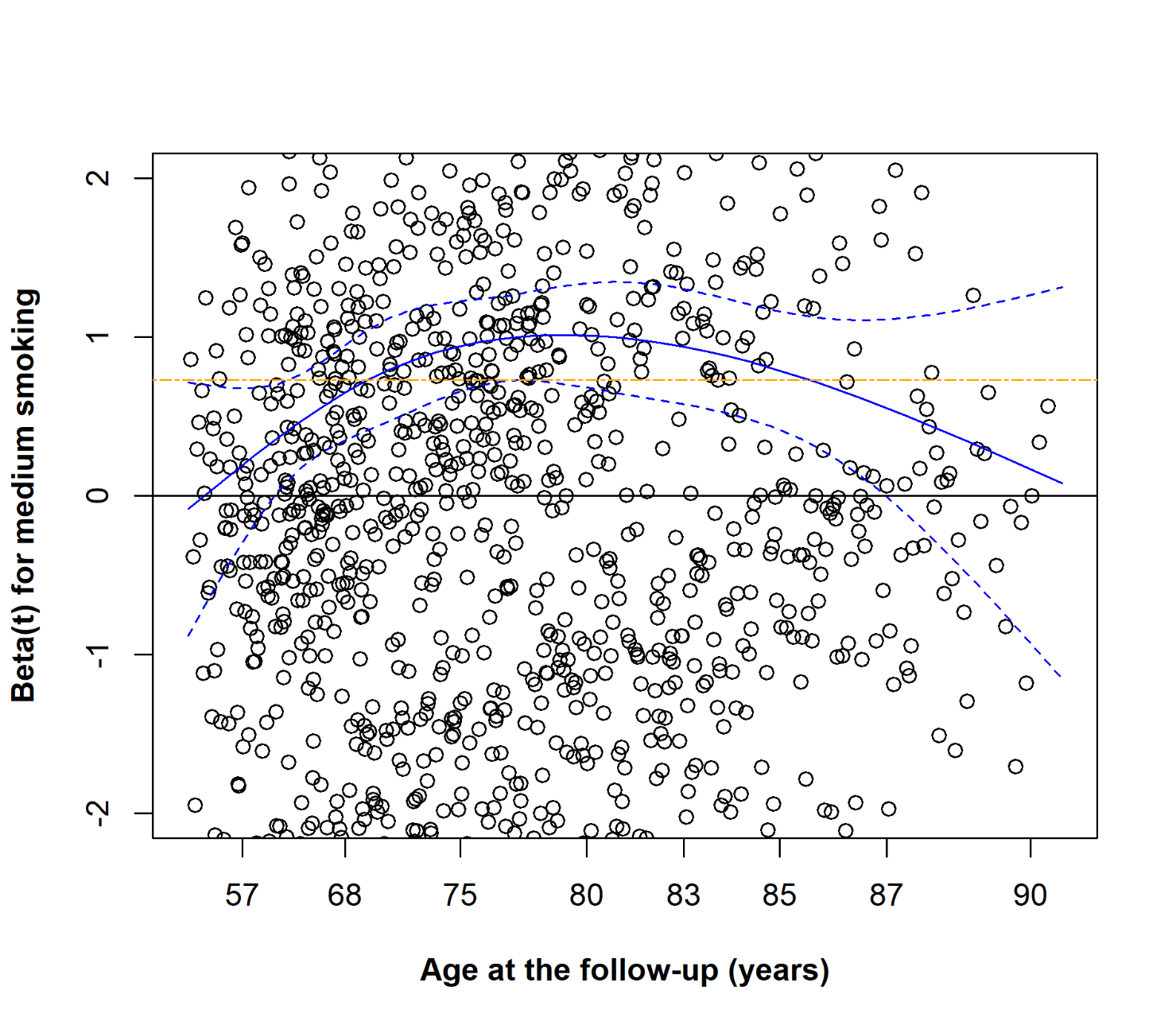

Figure S13: Schoenfeld residual plot of the association between the medium smoking group and the risk of all-cause mortality in the full model.

The association of the former smoking group with all-cause mortality was adjusted for the first ten genetic principal components of ancestry and family relatedness. The model also included the polygenic lifespan score, leisure-time physical activity, body mass index, alcohol consumption, occasional smoking group, former smoking group, light smoking group, heavy smoking group, and education level as covariates. The Schoenfeld residuals for each smoking behaviour group were obtained from models in which each group was recoded as a binary covariate (group vs. never smoking group). The solid black line is the reference line for zero residual deviation. The solid blue line indicates a smoothing spline of the fit and the dashed blue lines indicate a ±2-standard-error band around the fit. The two-dash orange line represents the *β_9_* reference line of heavy smoking in the model. Each circle represents a scaled Schoenfeld residual for an individual event (observation), illustrating the deviation from the proportional hazards assumption over time.

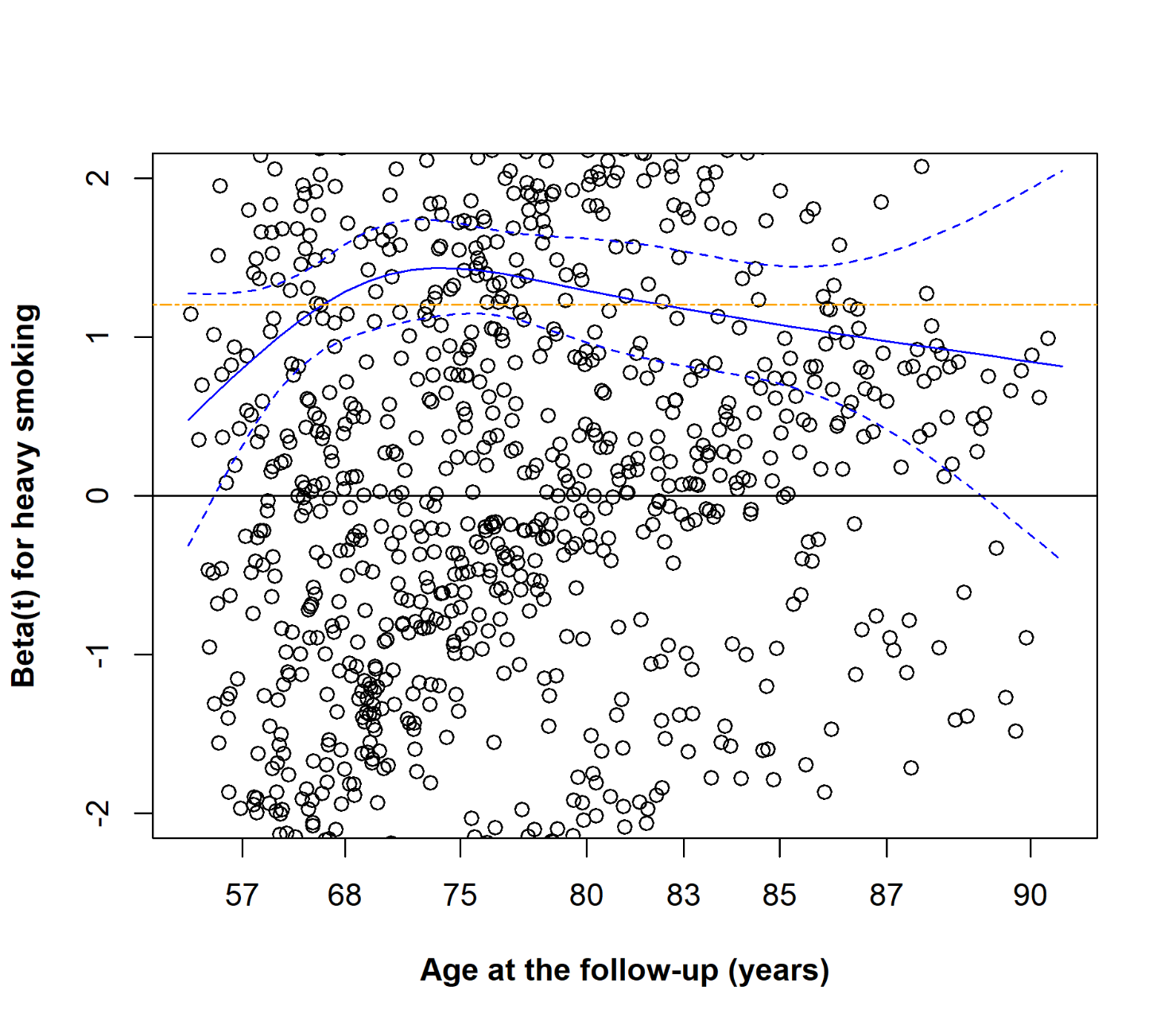

Figure S14: Schoenfeld residual plot of the association between the heavy smoking group and the risk of all-cause mortality in the full model.

The association of the former smoking group with all-cause mortality was adjusted for the first ten genetic principal components of ancestry and family relatedness. The model also included the polygenic lifespan score, leisure-time physical activity, body mass index, alcohol consumption, occasional smoking group, former smoking group, light smoking group, medium smoking group, and education level as covariates. The Schoenfeld residuals for each smoking behaviour group were obtained from models in which each group was recoded as a binary covariate (group vs. never smoking group). The solid black line is the reference line for zero residual deviation. The solid blue line indicates a smoothing spline of the fit and the dashed blue lines indicate a ±2-standard-error band around the fit. The two-dash orange line represents the *β*_10_ reference line of heavy smoking in the model. Each circle represents a scaled Schoenfeld residual for an individual event (observation), illustrating the deviation from the proportional hazards assumption over time.

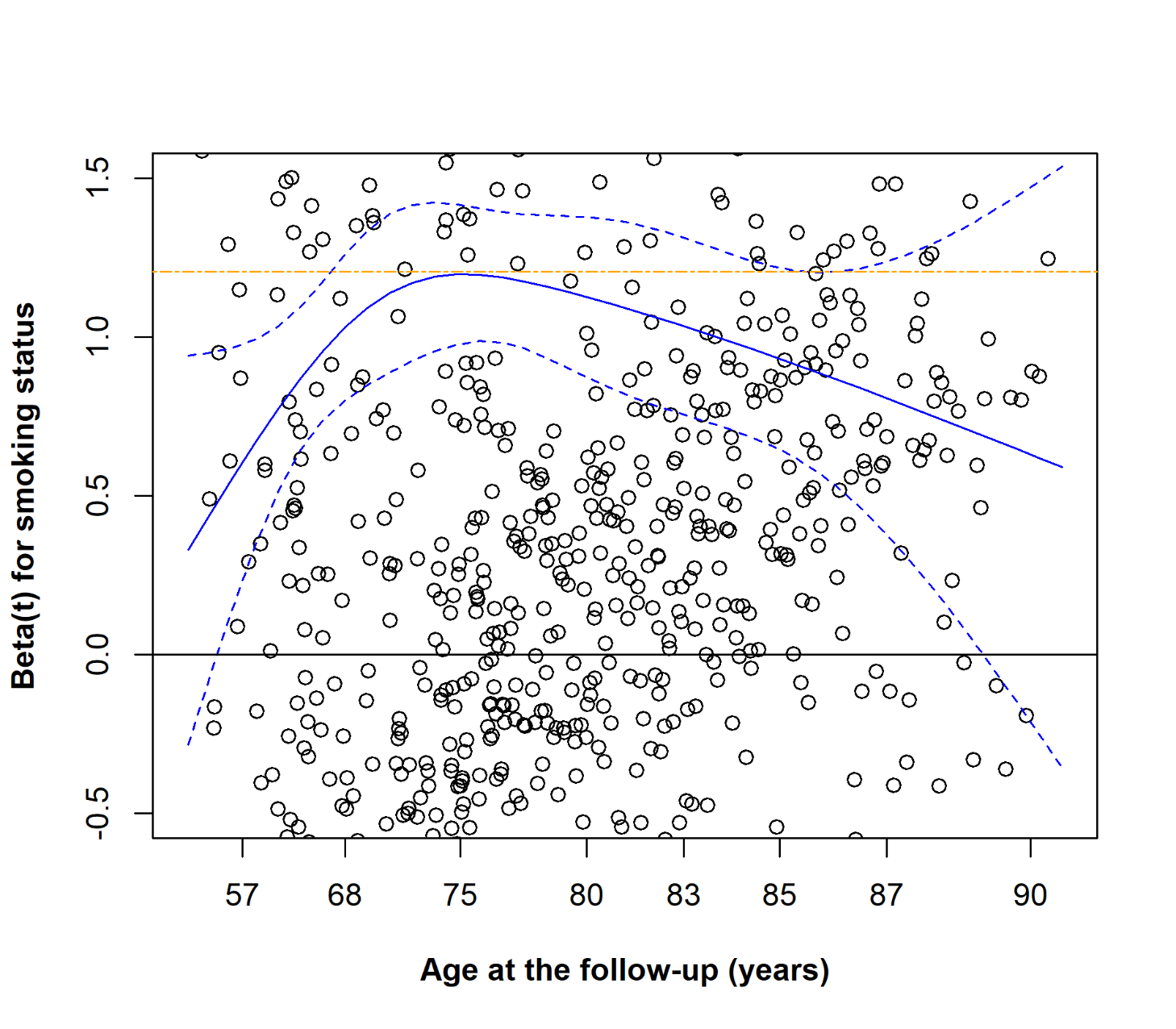

Figure S15: Schoenfeld residual plot of the association between all smoking behaviour groups and the risk of all-cause mortality in the full model.

The association of all smoking behaviour groups with all-cause mortality was adjusted for the first ten genetic principal components of ancestry and family relatedness. The model also included the polygenic lifespan score, leisure-time physical activity, body mass index, alcohol consumption, and education level as covariates. The solid black line is the reference line for zero residual deviation. The Schoenfeld residuals for this graph were obtained from the smoking behaviour covariate, which consists of all smoking behaviour groups (no binary smoking behaviour covariates). The solid blue line indicates a smoothing spline of the fit and the dashed blue lines indicate a ±2-standard-error band around the fit. The two-dash orange line represents the *β*_10_ reference line of heavy smoking (obtained from the binary heavy smoking group covariate) in the model. Each circle represents a scaled Schoenfeld residual for an individual event (observation), illustrating the deviation from the proportional hazards assumption over time.

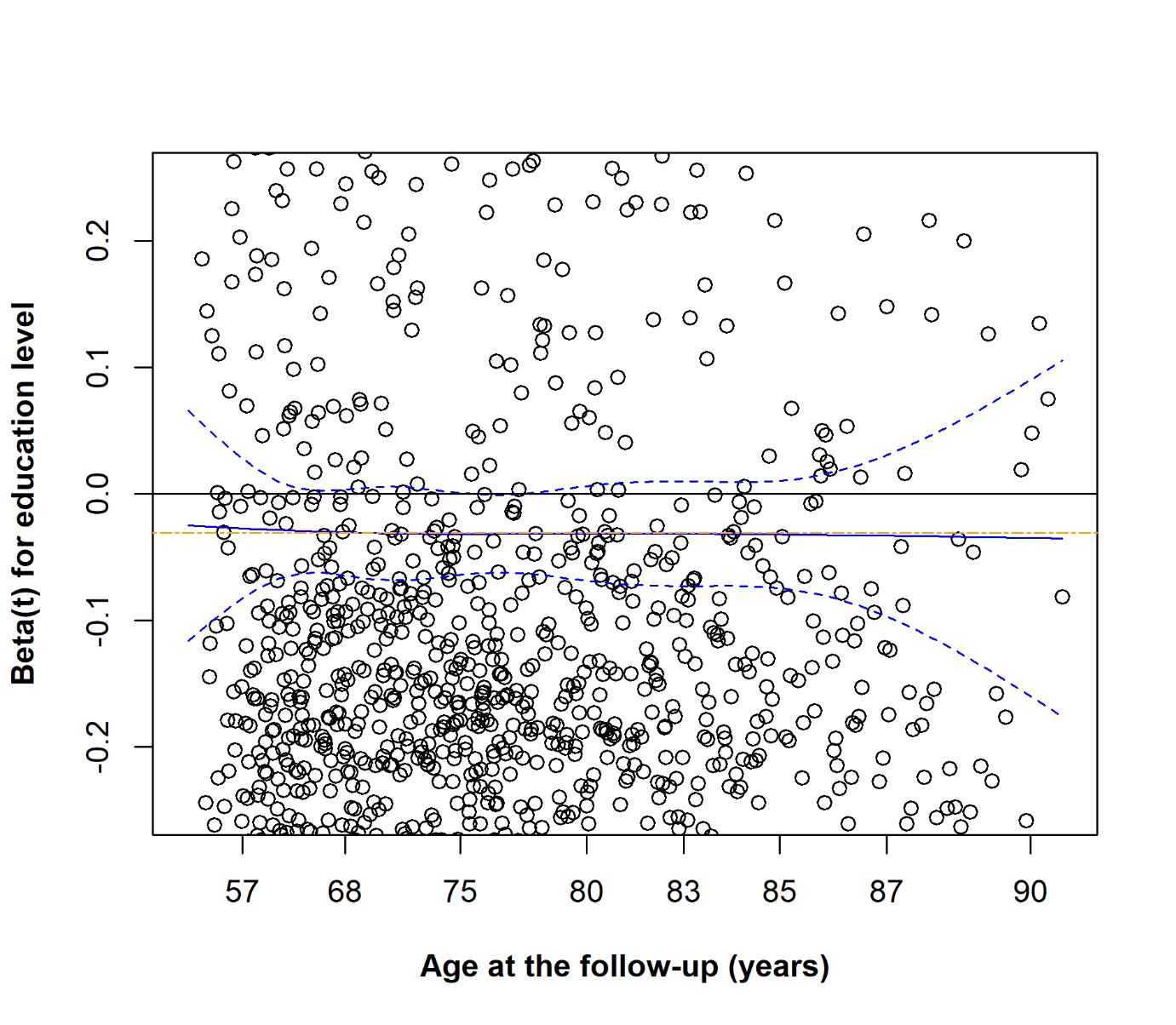

Figure S16: Schoenfeld residual plot of the association between education level and the risk of all-cause mortality in the full model.

The association of education level with all-cause mortality was adjusted for the first ten genetic principal components of ancestry and family relatedness. The model also included the polygenic lifespan score, leisure-time physical activity, body mass index, alcohol consumption, and smoking behaviour as covariates. The solid black line is the reference line for zero residual deviation. The solid blue line indicates a smoothing spline of the fit and the dashed blue lines indicate a ±2-standard-error band around the fit. The two-dash orange line represents the *β*_11_ reference line of education level in the model. Each circle represents a scaled Schoenfeld residual for an individual event (observation), illustrating the deviation from the proportional hazards assumption over time.
